# A systematic review and meta-analysis of school-based preventive interventions targeting e-cigarette use among adolescents

**DOI:** 10.1101/2023.12.19.23300263

**Authors:** Lauren A. Gardner, Amy-Leigh Rowe, Nicola C. Newton, Lyra Egan, Emily Hunter, Emma K. Devine, Tess Aitken, Louise Thornton, Maree Teesson, Emily Stockings, Katrina E. Champion

**Affiliations:** The Matilda Centre for Research in Mental Health and Substance Use, University of Sydney, Sydney, Australia; University of Sydney Library, University of Sydney, Sydney, Australia; School of Medicine and Public Health, The University of Newcastle, Newcastle, Australia

**Author notes:** Corresponding author: Dr. Lauren Gardner, The Matilda Centre for Research in Mental Health and Substance Use, University of Sydney, Sydney, Australia. Equally credited senior authors (Emily Stockings, Katrina E. Champion).

**Keywords:** E-cigarettes, vaping, school, intervention, prevention

## Abstract

**Objective:** To examine the efficacy of school-based e-cigarette preventive interventions via a systematic review and meta-analysis.

**Data sources:** We searched MEDLINE, Embase, PsycINFO, Scopus, CINAHL, Cochrane and clinical trials registries for studies published between January 2000-June 2023 using keywords for e-cigarettes, adolescents, and school.

**Study selection:** Of 1566 double-screened records, 11 met criteria of targeting adolescents, evaluating an e-cigarette preventive intervention, being conducted in a secondary school, using a randomised controlled trial (RCT), cluster RCT or quasi-experimental design, and comparing an intervention to a control.

**Data extraction:** Pre-specified data pertaining to the study design, outcomes, and quality were extracted by one reviewer and confirmed by a second, and where necessary, third reviewer.

**Data synthesis:** Our narrative synthesis showed some school-based interventions prevented or reduced e-cigarette and/or tobacco use, however some increased use. Meta-analyses on a subsample of studies found that, overall, school-based interventions were not associated with the prevention of e-cigarette (OR=0.43, 95%CI=0.16, 1.12; p=0.09) or tobacco (OR=1.01, 95%CI=0.65, 1.59, p=0.95) use, however were associated with reductions in past 30-day tobacco use (OR=0.59, 95%CI=0.39, 0.89, p=0.01) which encompassed e-cigarettes in some studies. School-based interventions were also associated with improved knowledge (SMD=-0.38, 95%CI=-0.68, -0.08, p=0.01), intentions (SMD=-0.15, 95%CI=-0.22, -0.07, p=0.0001), and attitudes (SMD=-0.14, 95%CI=-0.22, -0.06; p=0.0007) in the short-term. Overall, the quality of evidence was low-to-moderate.

**Conclusions:** School-based interventions hold potential for addressing e-cigarette use, however, can have null or iatrogenic effects. More high-quality research is needed to develop efficacious interventions, and schools must be supported to adopt evidence-based programs.

---

The growing use of electronic cigarettes (‘e-cigarettes’, also known as ‘vapes’) among young people is a critical public health concern due to the significant potential for harm. For example, e-cigarettes can cause a range of health issues including respiratory disease, seizures, poisoning, and injuries.^1^ E-cigarettes have also been associated with adverse mental health outcomes among young people including depression and suicidal ideation, with relatively little known about the longer-term health effects.^1^ ^2^ Additionally, adolescents are particularly susceptible to nicotine addiction, putting the developing brain at risk of damage.^1^ ^3^ In Australia, approximately one in four adolescents aged 14-17 have tried e-cigarettes, with one in ten reporting past 30-day use.^4^ Rates of past 30-day use are also high among adolescents in Canada and the United States, at 13% and 10%, respectively.^5^ ^6^ Among 11-18-year-olds in the United Kingdom, 40% have used e-cigarettes and approximately 5% use at least monthly.^7^ Evidence suggests that e-cigarette use is also associated with an increased likelihood of tobacco cigarette smoking.^8^ ^9^ Indeed, recent Australian data shows that tobacco smoking has increased among young people for the first time in recent decades,^10^ with increases similarly observed in Canada.^11^ This is highly concerning, given the substantial health, economic and societal burden of tobacco smoking.^8^ ^12^ Effective preventive interventions and public health strategies are therefore urgently needed to address e-cigarette use among adolescents.

While policy-level prevention initiatives can be effective and continue to evolve, they are unlikely to eliminate e-cigarette use completely. For example, there are minimum age of purchase regulations in the United Kingdom and United States (US), with some US states aiming to reduce access, supply and use via banning online sales and flavours. Australia adopts a prescription-only model, restricting sales of nicotine-containing e-cigarettes to those over age 18, with a prescription from their GP, which *should* only be available via pharmacies or online via the Therapeutic Goods Administration personal importation scheme.^13^ However, research indicates that most e-cigarettes used by Australian adolescents are illegally obtained and contain nicotine.^14^ ^15^ Further regulations for Australia were recently announced and are due to commence at the beginning of 2024, including a ban on all disposable vapes (regardless of nicotine content) and restrictions on flavours and packaging, leading to concerns over the development of a black market. Despite the varied measures, rates of e-cigarette use among young people in these regions remain unacceptably high, highlighting the need to provide critical education and resistance skills training to prevent or reduce the harms from e-cigarette use.

School-based interventions are an efficient, effective and economical prevention strategy. Schools offer an opportune setting to reach large numbers of adolescents during a critical period of exposure to substance use, and in an environment designed for learning and shaping of behaviour.^16^ ^17^ This is important as health habits formed during adolescence are likely to become entrenched.^18^ Further, adolescents’ social lives often revolve around the school context, meaning education can be delivered to peer groups whom can be strong influences on substance use behaviours.^16^ ^17^ Substance use preventive interventions are also feasible to implement in the school context, given drug education is typically mandatory within health education curriculums.^19^

Evidence shows that school-based interventions can be effective at preventing and/or delaying tobacco, alcohol and other substance use among adolescents,^17^ ^20^ ^21^ with some interventions demonstrating sustained effects into early adulthood.^22^ The strongest evidence exists for interventions that utilise a social competence and social influence approach to prevention.^23^ ^24^ As adolescent e-cigarette use and related harms have grown rapidly in recent years, so to have the number of interventions aiming to reduce these issues. However, to our knowledge, no study has systematically examined and meta-analysed the efficacy of school-based preventive interventions for e-cigarette use. This study therefore aims to:

1. Evaluate the efficacy of school-based preventive interventions in preventing e-cigarette use and improving secondary outcomes (e.g., tobacco use, knowledge, intentions, attitudes, and mental health) among adolescents.
2. Identify and summarise the key components and characteristics associated with efficacious interventions.

## Methods

### Approach and search terminology

This systematic review was conducted in accordance with the published review protocol^25^ and Preferred Reporting Items for Systematic Reviews and Meta-Analyses (PRISMA) guidelines^26^, and was prospectively registered with the International Prospective Register of Systematic Reviews (PROSPERO; CRD42022323352).

A research librarian (TA) conducted a systematic search of MEDLINE (Ovid), Embase (Ovid), PsycINFO (Ovid), Scopus, CINAHL, the Cochrane Database of Systematic Reviews (Ovid) and international clinical trial registries via the Cochrane Central Register of Controlled Trials (Ovid) for studies published from January 2000 (to slightly precede the advent of e-cigarettes in 2003 and thus ensure full capture of studies) to June 2022, which was re-run in June 2023. The search was limited to human studies, had no language restrictions, and was designed to identify grey literature and unpublished work, including dissertations, clinical trial registries and conference proceedings. An example search strategy is provided in Supplemental Table S1. All records identified in the search strategy were exported into EndNote and uploaded to the Covidence online software programme for deduplication and screening. Reference lists of eligible papers were reviewed to identify other relevant studies.

### Eligibility criteria

To be eligible for inclusion, studies must have: (1) targeted adolescents aged between 11 and 18 years of age at study intake (i.e., those of secondary school age); (2) evaluated an intervention targeting the prevention of e-cigarette use; (3) been conducted in a secondary school setting, however school-based interventions incorporating additional components (such as family-based or community-based elements) were also eligible; (4) used a randomised controlled trial, cluster randomised controlled trial, or a quasi-experimental design; and, (5) compared an intervention group to a control group (did not receive an intervention, education as usual, or alternative intervention). Interventions addressing other risk behaviours in addition to e-cigarette use (e.g., tobacco use or illicit drug use), and of both universal (i.e., delivered to all students in the intervention condition, regardless of their level of risk) and selective (i.e., delivered to higher-risk students in the intervention condition) nature were eligible.

### Exclusion criteria

Studies were excluded on the basis of: (1) not targeting adolescents; (2) not directly addressing e-cigarette use in the intervention; (3) the intervention having no school-based components; and, (4) having a non-experimental design or no control group.

### Article Screening and Coding

Two reviewers (LAG and AR) independently screened 1566 title and abstract records (98% agreement), as well as 36 full text records (94% agreement). Discrepancies at both screening stages were resolved by a third reviewer (KEC), resulting in 11 articles eligible for inclusion (see PRISMA flow diagram in Supplemental Figure S1). Data were extracted by one reviewer (EH or LE) and confirmed by a second (LAG), and where necessary, third (ES) reviewer. The following information from each eligible article was extracted: publication details, study characteristics, participant characteristics, intervention characteristics, primary and secondary outcomes of interest across all time points, measurement tools employed, details of the comparison group, data to assess risk of bias, and process data to determine the degree to which an intervention was implemented as intended. Corresponding authors were contacted for missing data. Further information on data extraction can be found in the published protocol.^25^

### Data analysis

The primary outcome was the prevention of e-cigarette use (lifetime use/ever use) at longest follow-up, treated as a dichotomous variable where N of participants in the intervention group reporting e-cigarette use is compared with N of participants in the control group reporting e-cigarette use. Raw Ns were extracted or reverse engineered, where possible, otherwise effect sizes and their confidence intervals were extracted. Secondary outcomes included the reduction or cessation of e-cigarette use (past 30-day) among adolescents already reporting e-cigarette use at baseline; the prevention (lifetime use/ever use) and reduction/cessation (past 30-day) of tobacco cigarette use; knowledge, attitudes (termed ‘outcome expectations’ in some studies), normative beliefs, harm perceptions, and future intentions related to e-cigarette/tobacco use; mental health outcomes; and, other substance use.

A narrative synthesis of study and intervention characteristics was conducted. When examining the characteristics associated with efficacious interventions, programs were deemed efficacious if a significant difference (at *p* < 0.05) was reported between the intervention and control groups on at least one primary or secondary outcome.

For the meta-analysis, data were entered into RevMan (version 6.7.1, Cochrane, London, UK) and combined using an inverse variance weighted random-effects analysis. Dichotomous outcomes are reported using odds ratios (ORs) and continuous outcomes using standardised mean difference (SMD). Where Standard Deviations were not reported, they were calculated. In line with recommendations in the Cochrane Handbook,^27^ where some scales increased for an outcome (e.g., higher scores indicate greater knowledge about e-cigarettes) whilst others decreased (e.g., lower scores indicate greater knowledge about e-cigarettes), the mean values from one set of studies was multiplied by -1 to ensure all scales pointed the same direction. This occurred for two secondary outcomes: knowledge and harm perceptions. Where improvement in an outcome is associated with higher scores on a scale, SMDs greater than zero indicate the degree to which the intervention is more efficacious than control, and SMDs less than zero indicate the degree to which treatment is less efficacious than placebo. Where improvement is associated with lower scores on a scale, SMDs lower than zero indicate the degree to which the intervention is more efficacious than control and SMDs greater than zero indicate the degree to which the intervention is less efficacious than control.^28^ If a study reported data at multiple timepoints, e-cigarette/tobacco use outcomes were meta-analysed at the longest follow-up timepoint, as per our prespecified primary outcome and given substance use behaviour change is more likely to be observed over the longer time period, as exposure increases throughout adolescence.^16^ ^21^ Conversely, outcomes theorised to precede behaviour change and exhibit change over the shorter-term (e.g., knowledge, attitudes, beliefs, and intentions) were examined at the first post-intervention timepoint.^29^ ^30^ Results are presented using forest plots and heterogeneity was assessed using the Higgins *I*^2^ statistic, with values of 25%, 50% and 75%, representing small, moderate or large heterogeneity, respectively. The significance of any heterogeneity was examined using the Cochran’s Q (χ2) test (p<0.05). Where appropriate, sub-analyses were conducted to explore sources of significant heterogeneity.

Two authors (AR and EKD [first search]; AR and LAG [updated search]) independently conducted the risk of bias assessment. Randomised studies were assessed using the Cochrane Collaboration’s tool for assessing risk of bias (RoB 2)^31^ and non-randomised studies were assessed using the Risk of Bias in Non-randomised Studies of Interventions (ROBINS-I).^32^ There was 100% agreement between all raters on the overall risk of bias for each of the included studies. The certainty of primary and secondary outcomes included in the meta-analysis was assessed using the Cochrane Grading of Recommendations Assessment, Development and Evaluation (GRADE) Framework.^33^

## Results

### Study characteristics

Of 1566 records identified, 36 full-text articles were screened, of which 11 were deemed eligible for inclusion. Nine studies provided sufficient data for meta-analysis.^34–42^ Table 1 provides characteristics of each study. Overall, the 11 articles included five cluster RCTs and six quasi-experimental trials. The studies comprised a total of 36,275 students, with sample sizes ranging from 158 to 13,269 students. Students were between 12 to 21 years at baseline and 50.98% were female. Trials were conducted between 2014-2020, most commonly in the US (n=5), and length of follow-up varied from immediately post-intervention to 36-months. Comparison groups included assessment only (n=6), education as usual (n=4), and an education-only component of an intervention (n=1). All studies used self-report survey measures to assess their outcomes. Baseline e-cigarette use, which sometimes encompassed broader tobacco use, was reported in 9 of the studies,^35^ ^36^ ^38–44^ of which six reported on lifetime use with prevalence rates ranging from 0%^44^ (due to only including never users at baseline) to 13.4%^41^. Three studies reported on past 30-day e-cigarette use at baseline, with prevalence rates of 2.2%,^40^ 4.3%^43^, and 36%^38^ (the latter being a combined measure of tobacco and e-cigarette use). Some studies (n=6) also measured other substance use at baseline, which included tobacco use (n=5), straw cigarettes (n=1), water pipe/ hookah (n=4), alcohol (n=2), marijuana (n=2), and hard drugs and crystal meth (n=1).

**Table 1.**
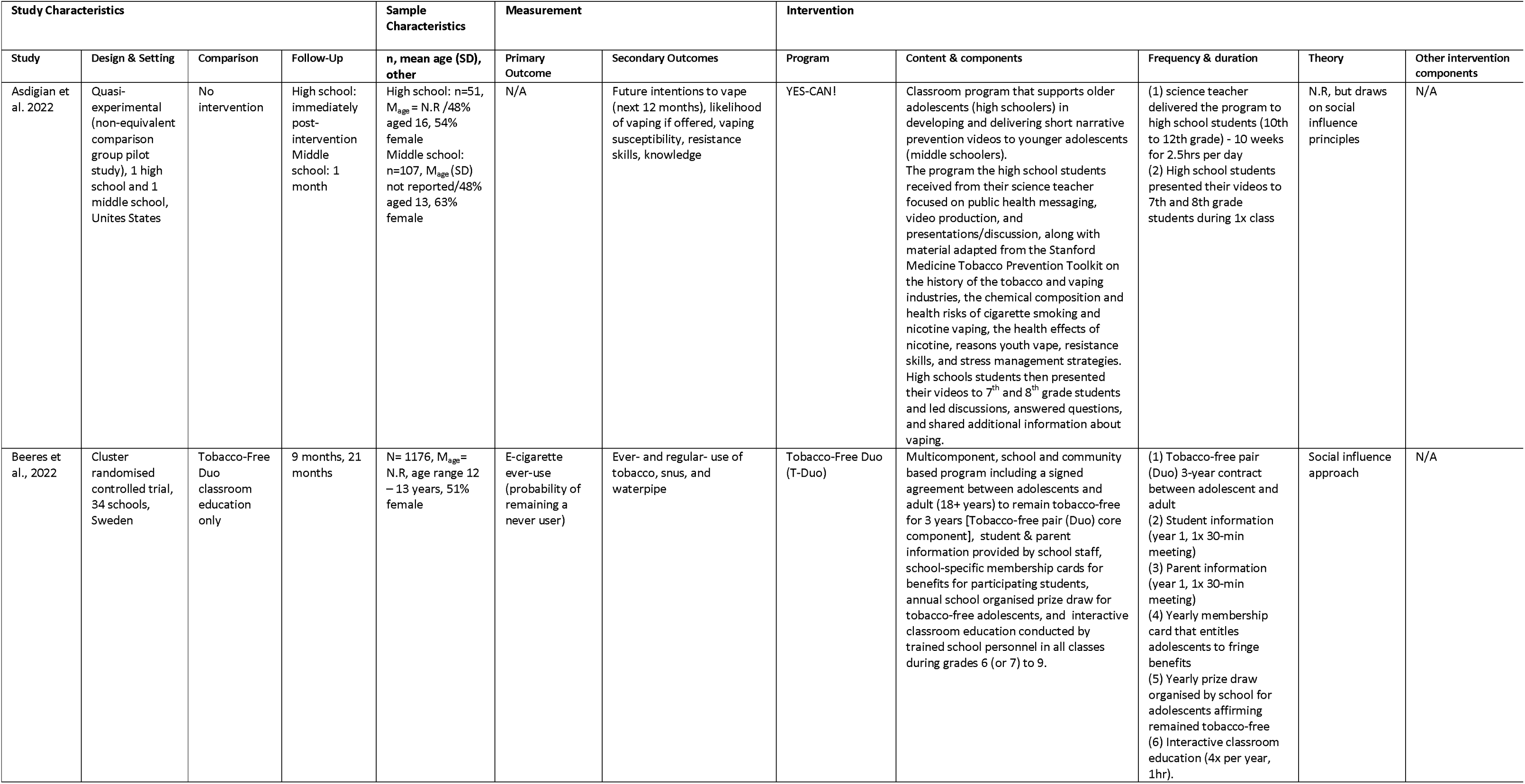
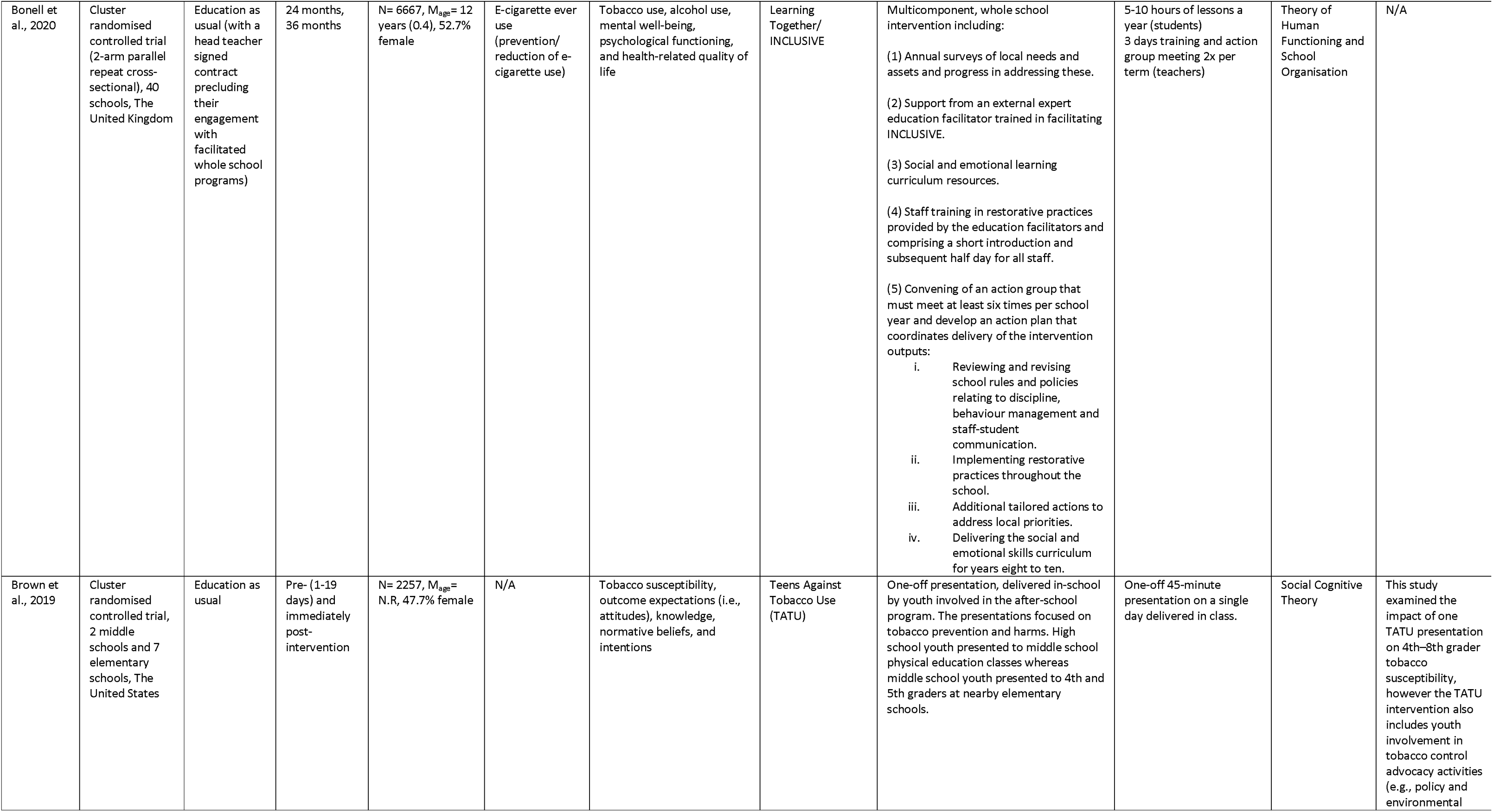
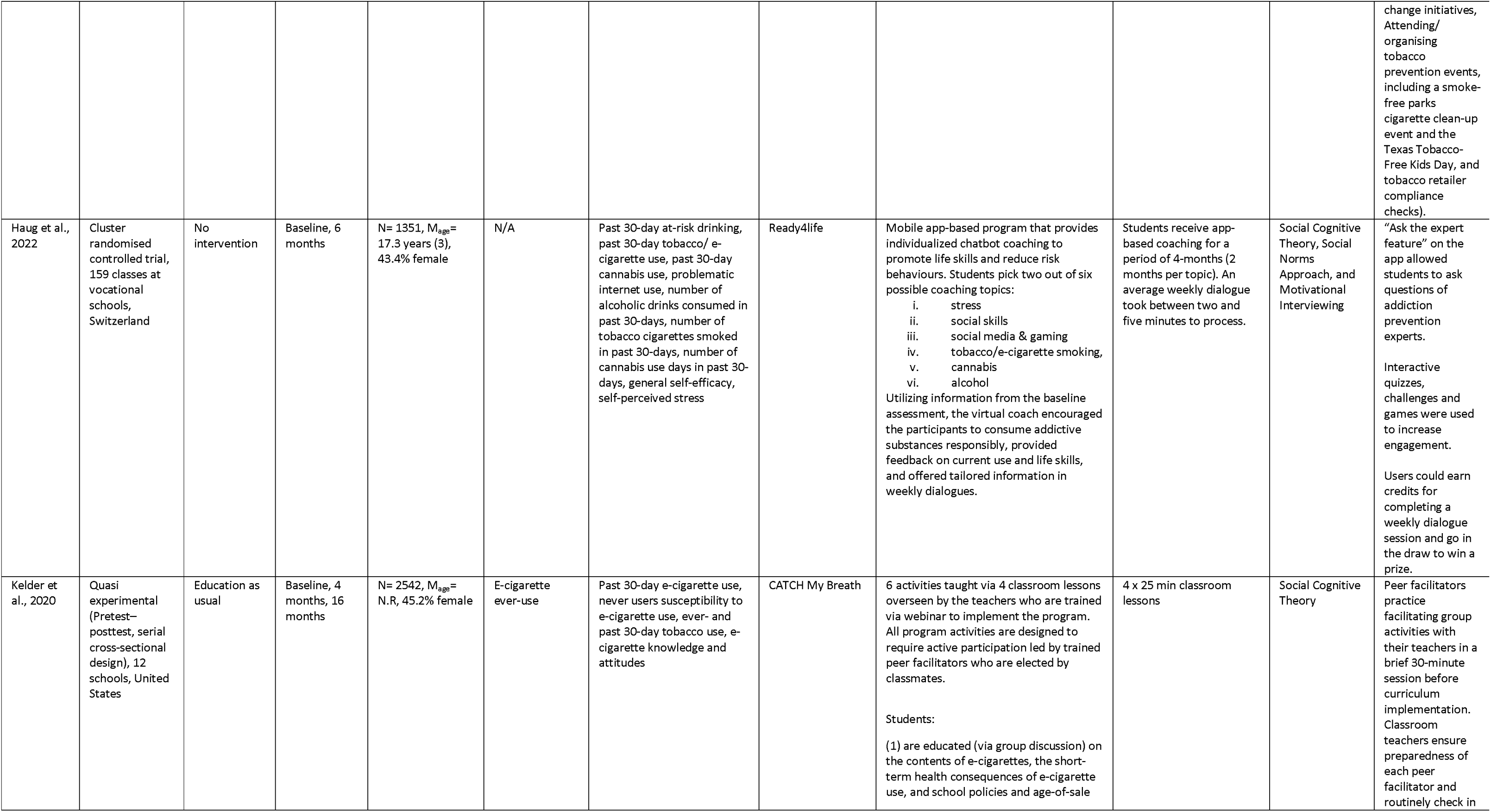
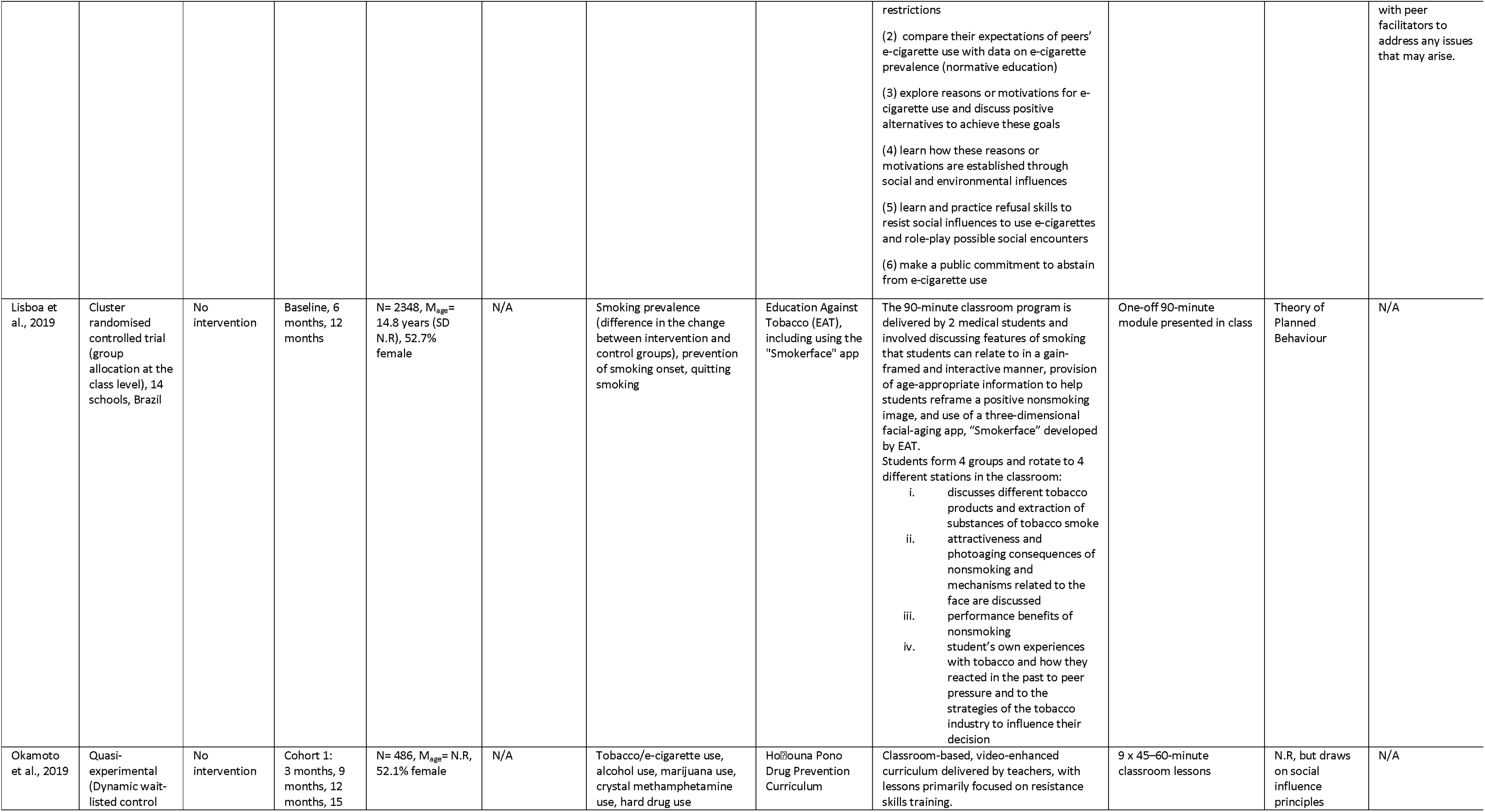
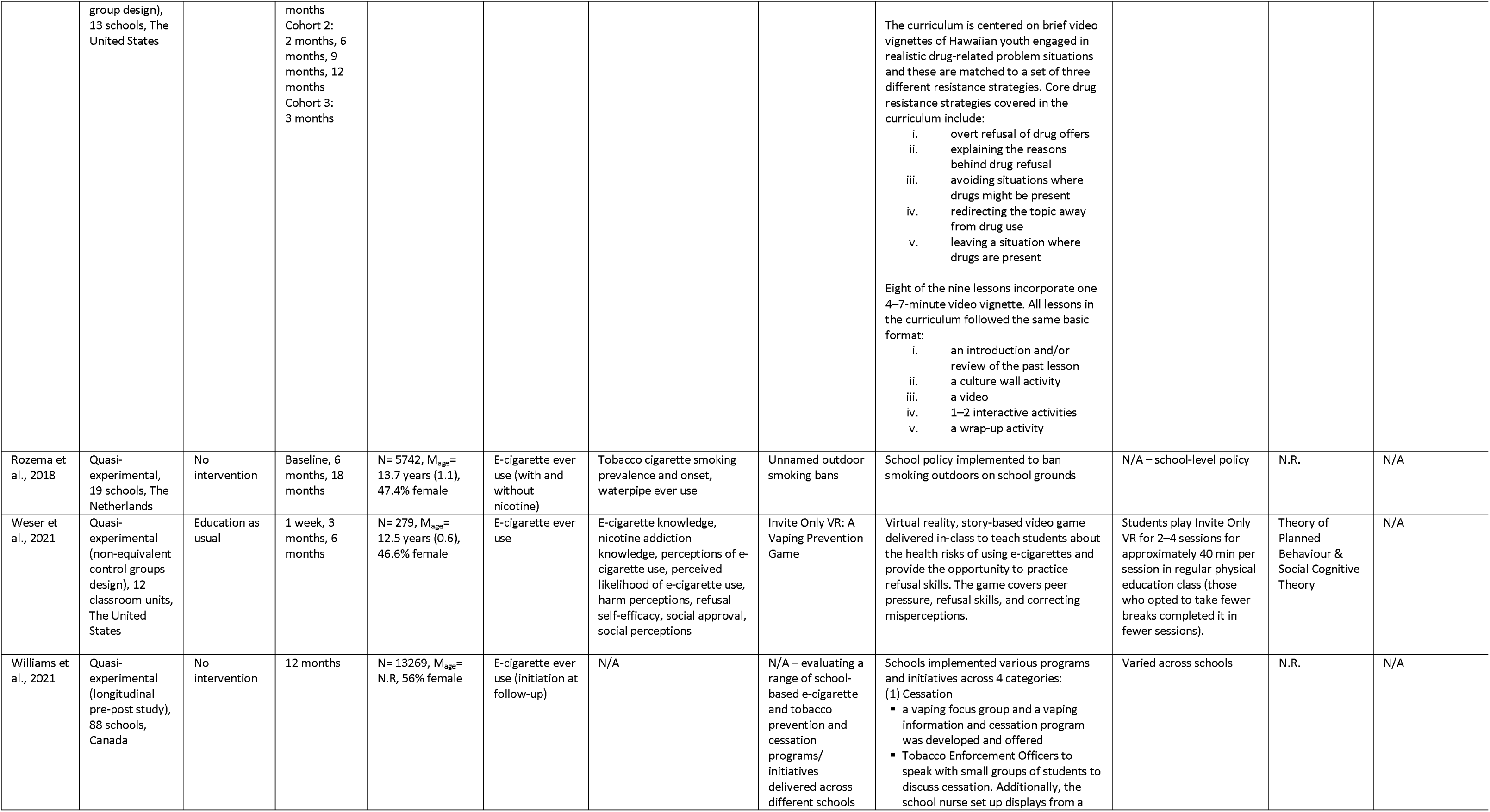
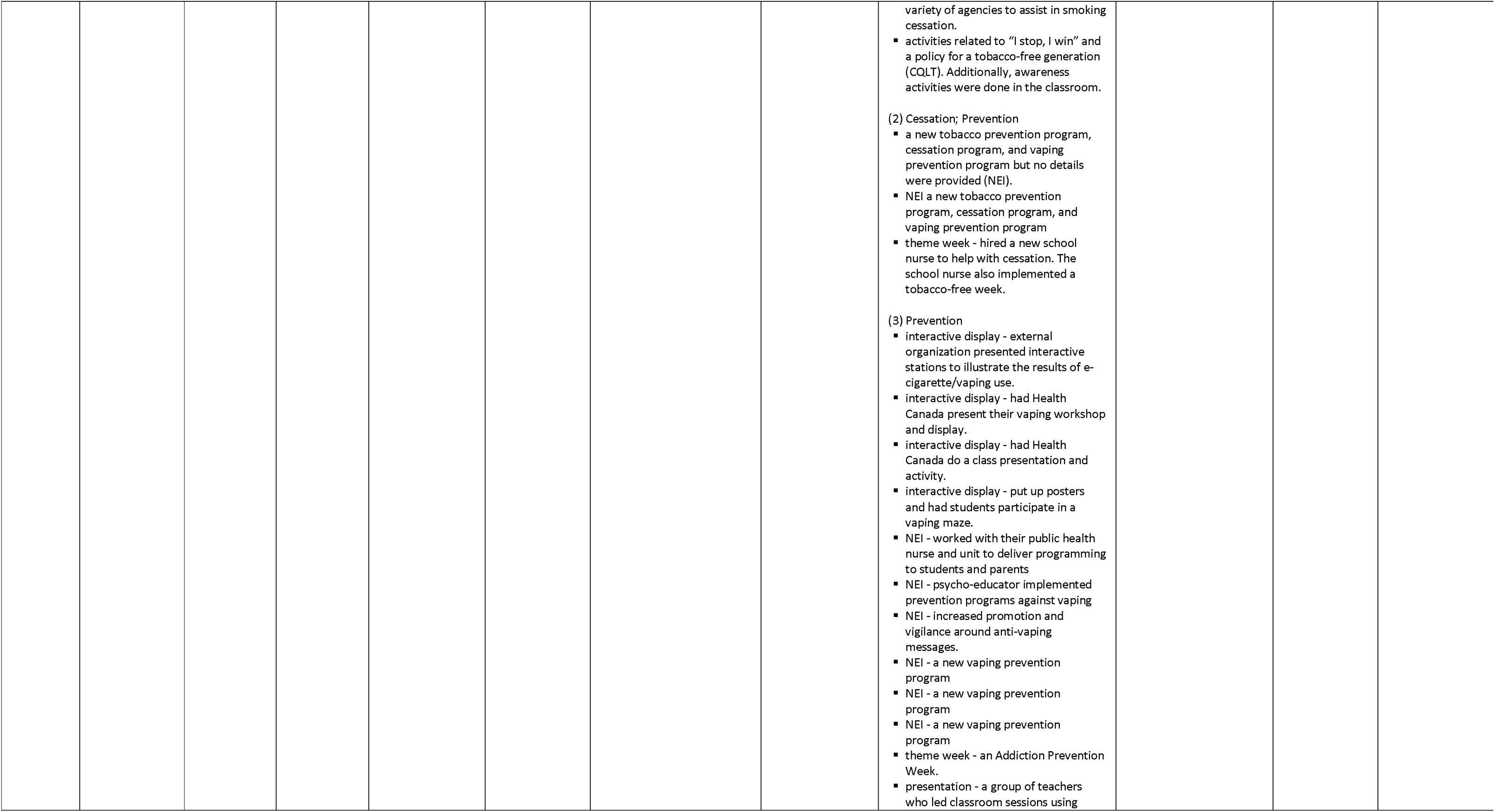
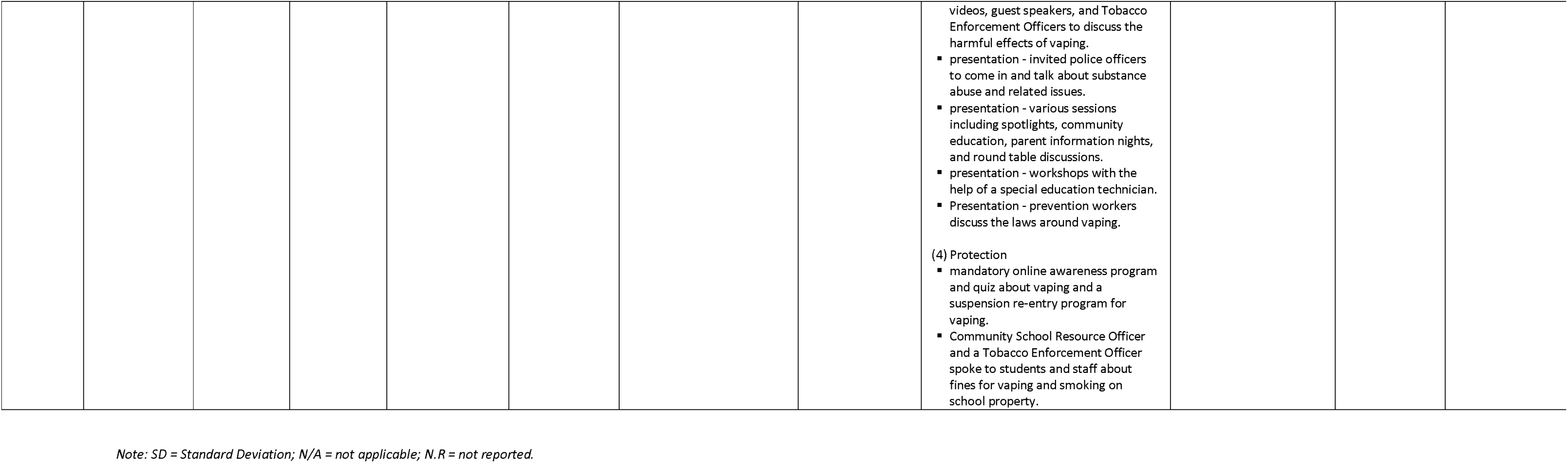
Characteristics of included studies.

| Study Characteristics |  |  |  | Sample Characteristics | Measurement |  | Intervention |  |  |  |  |
| --- | --- | --- | --- | --- | --- | --- | --- | --- | --- | --- | --- |
| Study | Design & Setting | Comparison | Follow-Up | n, mean age (SD), other | Primary Outcome | Secondary Outcomes | Program | Content & components | Frequency & duration | Theory | Other intervention components |
| Asdigian et al. 2022 | Quasi-experimental (non-equivalent comparison group pilot study), 1 high school and 1 middle school, Unites States | No intervention | High school: immediately post-intervention<br>Middle school: 1 month | High school: n=51, M<sub>age</sub> = N.R./48% aged 16, 54% female<br>Middle school: n=107, M<sub>age</sub> (SD) not reported/48% aged 13, 63% female | N/A | Future intentions to vape (next 12 months), likelihood of vaping if offered, vaping susceptibility, resistance skills, knowledge | YES-CAN! | Classroom program that supports older adolescents (high schoolers) in developing and delivering short narrative prevention videos to younger adolescents (middle schoolers).<br>The program the high school students received from their science teacher focused on public health messaging, video production, and presentations/discussion, along with material adapted from the Stanford Medicine Tobacco Prevention Toolkit on the history of the tobacco and vaping industries, the chemical composition and health risks of cigarette smoking and nicotine vaping, the health effects of nicotine, reasons youth vape, resistance skills, and stress management strategies. High schools students then presented their videos to 7<sup>th</sup> and 8<sup>th</sup> grade students and led discussions, answered questions, and shared additional information about vaping. | (1) science teacher delivered the program to high school students (10th to 12th grade) - 10 weeks for 2.5hrs per day<br>(2) High school students presented their videos to 7th and 8th grade students during 1x class | N.R, but draws on social influence principles | N/A |
| Beeres et al., 2022 | Cluster randomised controlled trial, 34 schools, Sweden | Tobacco-Free Duo classroom education only | 9 months, 21 months | N= 1176, M<sub>age</sub> = N.R, age range 12 –13 years, 51% female | E-cigarette ever-use (probability of remaining a never user) | Ever- and regular- use of tobacco, snus, and waterpipe | Tobacco-Free Duo (T-Duo) | Multicomponent, school and community based program including a signed agreement between adolescents and adult (18+ years) to remain tobacco-free for 3 years [Tobacco-free pair (Duo) core component], student & parent information provided by school staff, school-specific membership cards for benefits for participating students, annual school organised prize draw for tobacco-free adolescents, and interactive classroom education conducted by trained school personnel in all classes during grades 6 (or 7) to 9. | (1) Tobacco-free pair (Duo) 3-year contract between adolescent and adult<br>(2) Student information (year 1, 1x 30-min meeting)<br>(3) Parent information (year 1, 1x 30-min meeting)<br>(4) Yearly membership card that entitles adolescents to fringe benefits<br>(5) Yearly prize draw organised by school for adolescents affirming remained tobacco-free<br>(6) Interactive classroom education (4x per year, 1hr). | Social influence approach | N/A |
| Bonell et al., 2020 | Cluster randomised controlled trial (2-arm parallel repeat cross-sectional), 40 schools, The United Kingdom | Education as usual (with a head teacher signed contract precluding their engagement with facilitated whole school programs) | 24 months, 36 months | N= 6667, M<sub>age</sub> = 12 years (0.4), 52.7% female | E-cigarette ever use (prevention/reduction of e-cigarette use) | Tobacco use, alcohol use, mental well-being, psychological functioning, and health-related quality of life | Learning Together/ INCLUSIVE | Multicomponent, whole school intervention including:<br>(1) Annual surveys of local needs and assets and progress in addressing these.<br>(2) Support from an external expert education facilitator trained in facilitating INCLUSIVE.<br>(3) Social and emotional learning curriculum resources.<br>(4) Staff training in restorative practices provided by the education facilitators and comprising a short introduction and subsequent half day for all staff.<br>(5) Convening of an action group that must meet at least six times per school year and develop an action plan that coordinates delivery of the intervention outputs:<br>Reviewing and revising school rules and policies relating to discipline, behaviour management and staff-student communication.<br>Implementing restorative practices throughout the school.<br>Additional tailored actions to address local priorities.<br>Delivering the social and emotional skills curriculum for years eight to ten. | 5-10 hours of lessons a year (students)<br>3 days training and action group meeting 2x per term (teachers) | Theory of Human Functioning and School Organisation | N/A |
| Brown et al., 2019 | Cluster randomised controlled trial, 2 middle schools and 7 elementary schools, The United States | Education as usual | Pre- (1-19 days) and immediately post-intervention | N= 2257, M<sub>age</sub> = N.R, 47.7% female | N/A | Tobacco susceptibility, outcome expectations (i.e., attitudes), knowledge, normative beliefs, and intentions | Teens Against Tobacco Use (TATU) | One-off presentation, delivered in-school by youth involved in the after-school program. The presentations focused on tobacco prevention and harms. High school youth presented to middle school physical education classes whereas middle school youth presented to 4th and 5th graders at nearby elementary schools. | One-off 45-minute presentation on a single day delivered in class. | Social Cognitive Theory | This study examined the impact of one TATU presentation on 4th–8th grader tobacco susceptibility, however the TATU intervention also includes youth involvement in tobacco control advocacy activities (e.g., policy and environmental change initiatives, Attending/ organising tobacco prevention events, including a smoke-free parks cigarette clean-up event and the Texas Tobacco-Free Kids Day, and tobacco retailer compliance checks). |
| Haug et al., 2022 | Cluster randomised controlled trial, 159 classes at vocational schools, Switzerland | No intervention | Baseline, 6 months | N= 1351, M<sub>age</sub> = 17.3 years (3), 43.4% female | N/A | Past 30-day at-risk drinking, past 30-day tobacco/ e-cigarette use, past 30-day cannabis use, problematic internet use, number of alcoholic drinks consumed in past 30-days, number of tobacco cigarettes smoked in past 30-days, number of cannabis use days in past 30-days, general self-efficacy, self-perceived stress | Ready4life | Mobile app-based program that provides individualized chatbot coaching to promote life skills and reduce risk behaviours. Students pick two out of six possible coaching topics:<br>i. stress<br>ii. social skills<br>iii. social media & gaming<br>iv. tobacco/e-cigarette smoking,<br>v. cannabis<br>vi. alcohol<br>Utilizing information from the baseline assessment, the virtual coach encouraged the participants to consume addictive substances responsibly, provided feedback on current use and life skills, and offered tailored information in weekly dialogues. | Students receive app-based coaching for a period of 4-months (2 months per topic). An average weekly dialogue took between two and five minutes to process. | Social Cognitive Theory, Social Norms Approach, and Motivational Interviewing | “Ask the expert feature” on the app allowed students to ask questions of addiction prevention experts.<br>Interactive quizzes, challenges and games were used to increase engagement.<br>Users could earn credits for completing a weekly dialogue session and go in the draw to win a prize. |
| Kelder et al., 2020 | Quasi experimental (Pretest–posttest, serial cross-sectional design), 12 schools, United States | Education as usual | Baseline, 4 months, 16 months | N= 2542, M<sub>age</sub> = N.R, 45.2% female | E-cigarette ever-use | Past 30-day e-cigarette use, never users susceptibility to e-cigarette use, ever- and past 30-day tobacco use, e-cigarette knowledge and attitudes | CATCH My Breath | 6 activities taught via 4 classroom lessons overseen by the teachers who are trained via webinar to implement the program. All program activities are designed to require active participation led by trained peer facilitators who are elected by classmates.<br>Students:<br>(1) are educated (via group discussion) on the contents of e-cigarettes, the short-term health consequences of e-cigarette use, and school policies and age-of-sale<br>restrictions<br>(2) compare their expectations of peers' e-cigarette use with data on e-cigarette prevalence (normative education)<br>(3) explore reasons or motivations for e-cigarette use and discuss positive alternatives to achieve these goals<br>(4) learn how these reasons or motivations are established through social and environmental influences<br>(5) learn and practice refusal skills to resist social influences to use e-cigarettes and role-play possible social encounters<br>(6) make a public commitment to abstain from e-cigarette use | 4 x 25 min classroom lessons | Social Cognitive Theory | Peer facilitators practice facilitating group activities with their teachers in a brief 30-minute session before curriculum implementation. Classroom teachers ensure preparedness of each peer facilitator and routinely check in with peer facilitators to address any issues that may arise. |
| Lisboa et al., 2019 | Cluster randomised controlled trial (group allocation at the class level), 14 schools, Brazil | No intervention | Baseline, 6 months, 12 months | N= 2348, M<sub>age</sub> = 14.8 years (SD N.R), 52.7% female | N/A | Smoking prevalence (difference in the change between intervention and control groups), prevention of smoking onset, quitting smoking | Education Against Tobacco (EAT), including using the "Smokerface" app | The 90-minute classroom program is delivered by 2 medical students and involved discussing features of smoking that students can relate to in a gain-framed and interactive manner, provision of age-appropriate information to help students reframe a positive nonsmoking image, and use of a three-dimensional facial-aging app, "Smokerface" developed by EAT.<br>Students form 4 groups and rotate to 4 different stations in the classroom:<br>i. discusses different tobacco products and extraction of substances of tobacco smoke<br>ii. attractiveness and photoaging consequences of nonsmoking and mechanisms related to the face are discussed<br>iii. performance benefits of nonsmoking<br>iv. student's own experiences with tobacco and how they reacted in the past to peer pressure and to the strategies of the tobacco industry to influence their decision | One-off 90-minute module presented in class | Theory of Planned Behaviour | N/A |
| Okamoto et al., 2019 | Quasi-experimental (Dynamic wait-listed control | No intervention | Cohort 1: 3 months, 9 months, 12 months, 15 | N= 486, M<sub>age</sub> = N.R, 52.1% female | N/A | Tobacco/e-cigarette use, alcohol use, marijuana use, crystal methamphetamine use, hard drug use | Holġouna Pono Drug Prevention Curriculum | Classroom-based, video-enhanced curriculum delivered by teachers, with lessons primarily focused on resistance skills training. | 9 x 45–60-minute classroom lessons | N.R, but draws on social influence principles | N/A |
|  | group design), 13 schools, The United States |  | months<br>Cohort 2:<br>2 months, 6 months, 9 months, 12 months<br>Cohort 3:<br>3 months |  |  |  |  | The curriculum is centered on brief video vignettes of Hawaiian youth engaged in realistic drug-related problem situations and these are matched to a set of three different resistance strategies. Core drug resistance strategies covered in the curriculum include:<br>overt refusal of drug offers<br>explaining the reasons behind drug refusal<br>avoiding situations where drugs might be present<br>redirecting the topic away from drug use<br>leaving a situation where drugs are present<br>Eight of the nine lessons incorporate one 4–7-minute video vignette. All lessons in the curriculum followed the same basic format:<br>an introduction and/or review of the past lesson<br>a culture wall activity<br>a video<br>1–2 interactive activities<br>a wrap-up activity |  |  |  |
| Rozema et al., 2018 | Quasi-experimental, 19 schools, The Netherlands | No intervention | Baseline, 6 months, 18 months | N= 5742, M<sub>age</sub>= 13.7 years (1.1), 47.4% female | E-cigarette ever use (with and without nicotine) | Tobacco cigarette smoking prevalence and onset, waterpipe ever use | Unnamed outdoor smoking bans | School policy implemented to ban smoking outdoors on school grounds | N/A – school-level policy | N.R. | N/A |
| Weser et al., 2021 | Quasi-experimental (non-equivalent control groups design), 12 classroom units, The United States | Education as usual | 1 week, 3 months, 6 months | N= 279, M<sub>age</sub>= 12.5 years (0.6), 46.6% female | E-cigarette ever use | E-cigarette knowledge, nicotine addiction knowledge, perceptions of e-cigarette use, perceived likelihood of e-cigarette use, harm perceptions, refusal self-efficacy, social approval, social perceptions | Invite Only VR: A Vaping Prevention Game | Virtual reality, story-based video game delivered in-class to teach students about the health risks of using e-cigarettes and provide the opportunity to practice refusal skills. The game covers peer pressure, refusal skills, and correcting misperceptions. | Students play Invite Only VR for 2–4 sessions for approximately 40 min per session in regular physical education class (those who opted to take fewer breaks completed it in fewer sessions). | Theory of Planned Behaviour & Social Cognitive Theory | N/A |
| Williams et al., 2021 | Quasi-experimental (longitudinal pre-post study), 88 schools, Canada | No intervention | 12 months | N= 13269, M<sub>age</sub>= N.R, 56% female | E-cigarette ever use (initiation at follow-up) | N/A | N/A – evaluating a range of school-based e-cigarette and tobacco prevention and cessation programs/ initiatives delivered across different schools | Schools implemented various programs and initiatives across 4 categories:<br>(1) Cessation<br>a vaping focus group and a vaping information and cessation program was developed and offered<br>Tobacco Enforcement Officers to speak with small groups of students to discuss cessation. Additionally, the school nurse set up displays from a | Varied across schools | N.R. | N/A |

|  |  |  |  |  |  |  |  |  |
| --- | --- | --- | --- | --- | --- | --- | --- | --- |
|  |  |  |  |  |  |  |  | variety of agencies to assist in smoking cessation.<br>▪ activities related to “I stop, I win” and a policy for a tobacco-free generation (CQLT). Additionally, awareness activities were done in the classroom.<br>(2) Cessation; Prevention<br>▪ a new tobacco prevention program, cessation program, and vaping prevention program but no details were provided (NEI).<br>▪ NEI a new tobacco prevention program, cessation program, and vaping prevention program<br>▪ theme week - hired a new school nurse to help with cessation. The school nurse also implemented a tobacco-free week.<br>(3) Prevention<br>▪ interactive display - external organization presented interactive stations to illustrate the results of e-cigarette/vaping use.<br>▪ interactive display - had Health Canada present their vaping workshop and display.<br>▪ interactive display - had Health Canada do a class presentation and activity.<br>▪ interactive display - put up posters and had students participate in a vaping maze.<br>▪ NEI - worked with their public health nurse and unit to deliver programming to students and parents<br>▪ NEI - psycho-educator implemented prevention programs against vaping<br>▪ NEI - increased promotion and vigilance around anti-vaping messages.<br>▪ NEI - a new vaping prevention program<br>▪ NEI - a new vaping prevention program<br>▪ NEI - a new vaping prevention program<br>▪ theme week - an Addiction Prevention Week.<br>▪ presentation - a group of teachers who led classroom sessions using |

|  |  |  |  |  |  |  |  |  |
| --- | --- | --- | --- | --- | --- | --- | --- | --- |
|  |  |  |  |  |  |  |  | videos, guest speakers, and Tobacco Enforcement Officers to discuss the harmful effects of vaping.<br>▪ presentation - invited police officers to come in and talk about substance abuse and related issues.<br>▪ presentation - various sessions including spotlights, community education, parent information nights, and round table discussions.<br>▪ presentation - workshops with the help of a special education technician.<br>▪ Presentation - prevention workers discuss the laws around vaping.<br>(4) Protection<br>▪ mandatory online awareness program and quiz about vaping and a suspension re-entry program for vaping.<br>▪ Community School Resource Officer and a Tobacco Enforcement Officer spoke to students and staff about fines for vaping and smoking on school property. |

*Note: SD = Standard Deviation; N/A = not applicable; N.R = not reported.*

### Intervention characteristics

Most interventions targeted e-cigarette and tobacco use concurrently (n=8^34^ ^35^ ^37–41^ ^43^), however one^42^ targeted e-cigarette use alone, and another^36^ focused more broadly on transforming the school environment through restorative approaches to address conflict, behaviour management policies and social and emotional learning. Additionally, in one study,^44^ multiple prevention and cessation programs and policy interventions were evaluated across 24 schools. Although the descriptions were limited, the interventions appeared to vary substantially (e.g., some focused on e-cigarettes and others included tobacco more broadly, some involved teacher-led sessions and others involved external facilitators, police officers or school nurses, some involved classroom education and others involved presentations, eHealth components, school policy, or theme weeks). Among the other eligible studies, interventions included in-class student information and skills training programs led by peers and/or teachers (n=6^34^ ^37^ ^39^ ^40^ ^42^ ^43^), multicomponent programs (n=2^35^ ^36^), school-wide policy (n=1^41^), and a mobile phone app (n=1^38^). Delivery methods comprised face-to-face (n=2^36^ ^37^), eHealth-only (n=2^38^ ^42^), hybrid (n=5^34^ ^35^ ^39^ ^40^ ^43^) and policy-only (n=1^41^) formats. All studies evaluated universal preventive interventions, however the one study that evaluated multiple programs across schools^44^ also included cessation programs for students already using e-cigarettes or tobacco cigarettes. Intervention duration and frequency ranged from a one-off 45-minute presentation^37^ to a 3-year tobacco-free contract between adolescents and an adult.^35^ Most interventions (n=9) were underpinned by behavioural theory, spanning: social cognitive theory (n=4); a social influence approach (n=3), the theory of planned behaviour (n=2); the theory of human functioning and school organisation (n=1); the social norms approach (n=1); and/or, motivational interviewing (n=1).

### Characteristics of efficacious interventions

Of the 11 trials, nine^34–40^ ^42^ ^43^ found a significant improvement on at least one primary or secondary outcome. Six studies reported specifically on the prevention of e-cigarette use,^35,36^ ^39^ ^41^ ^42^ ^44^ of which half found significant intervention effects in the expected direction.^35^ ^36^ ^39^ Two^35^ ^36^ of these three efficacious interventions were multicomponent programs, consisting of school-based components (e.g., classroom education and teacher training) along with broader initiatives involving parents or external adults. The other^39^ involved classroom education and skills training alone, including both teacher- and peer-led components. The classroom education in all three efficacious interventions was delivered over multiple sessions, ranging between 4x25min lessons to a 10hr learning curriculum, and was overseen by trained teachers. However, only two interventions^35^ ^39^ focused specifically on e-cigarettes/tobacco, both via a hybrid delivery format, whereas the other^36^ focused on broader behaviour management via social and emotional learning in a face-to-face format.

Of the seven trials that examined change in tobacco use – the definition of which varied across studies (e.g., referred to tobacco cigarettes specifically in some studies^35^ ^36^ ^41^ and more encompassed e-cigarettes and/or other types such as cigars, pipes, and hookah in other studies^38–40^ ^43^) – five reported significant intervention effects in the expected direction. Three of these were the same efficacious interventions that were associated with the prevention of e-cigarette use,^35^ ^36^ ^39^ with the addition of two other classroom education and skills training programs.^40^ One of which^40^ involved a one-off 90-minute peer-led module delivered in a hybrid format (face-to-face by medical students along with an eHealth component) and the other^43^ involved a nine-lesson, video-enhanced curriculum delivered by teachers. Theoretical underpinnings of these studies included the social cognitive theory (n=2^35^ ^39^), the theory of planned behaviour (n=1^40^), the theory of human functioning and school organisation (n=1^36^) and, although not explicitly stated, social influence principles (n=1^43^).

Iatrogenic effects on the prevention of e-cigarette use^44^ and tobacco smoking onset^41^ were reported within two studies. The two interventions included a broad “theme week”^44^ and a school-wide policy.^41^ Descriptions were limited, however neither of these interventions appeared to include classroom education and skills training or be grounded in theory.

Four studies were associated with a significant improvement in theoretical constructs related to e-cigarette and/or tobacco cigarette use, including knowledge,^34^ ^37^ ^39^ ^42^ attitudes,^34^ ^37^ ^39^ normative beliefs,^34^ harm perceptions,^42^ and future intentions to use tobacco.^37^ All of these interventions centred on classroom education and skills training, however some involved teacher- and peer-led components delivered over multiple lessons via hybrid format,^34^ ^39^ one involved a one-off peer-led face-to-face presentation,^37^ and the other comprised an eHealth program used over multiple lessons.^42^ The most common theoretical underpinning was the social cognitive theory (n=3^37^ ^39^ ^42^), with two interventions also drawing on social influence principles (n=1^34^) and the theory of planned behaviour (n=1^42^).

In terms of other secondary outcomes, two studies were associated with significantly better mental health outcomes, including lower perceived stress^38^ and greater mental wellbeing, psychological functioning, and health-related quality of life.^36^ Although both studies incorporated student education and skills training over multiple sessions, one did so via face-to-face format within a broader multicomponent intervention that additionally comprised staff training and school policies and practices,^36^ while the other used an eHealth intervention delivered outside of class.^38^ Both of these interventions were also associated with reduced alcohol use. Finally, two interventions were associated with the prevention or reduction of illicit drug use, including the broader multicomponent intervention focusing on social and emotional learning,^36^ and another intervention similarly involving student education and skills training over multiple sessions, however delivery was via a video-enhanced drug prevention curriculum.^43^ There were no clear characteristics of successful studies in terms of theoretical underpinnings.

Results from the meta-analyses of primary and secondary outcomes are presented below.

### Outcomes

#### Primary outcome: The prevention of e-cigarette use at longest follow-up

Five studies included primary outcome data sufficient for meta-analysis (see Figure 1). Overall, these studies found that school-based interventions were not significantly associated with the prevention of e-cigarette use at longest follow-up (OR=0.43, 95% CI=0.16, 1.12; p=0.09) and there was considerable heterogeneity between studies (I^2=^98%, p<0.00001). To explore sources of heterogeneity, we conducted sub-group analysis by intervention type (Figure 1). Interventions centring on student education and resistance skills training alone were not significantly associated with the prevention of e-cigarette use (OR=0.77, 95% CI=0.21, 2.86, p=0.69; I² = 45%, p=0.18). There was some evidence that broader school-based interventions prevented e-cigarette use, however this did not reach traditional significance levels (OR=0.31, 95% CI=0.08, 1.16, p=0.08) and substantial heterogeneity remained (I² = 99%, p< 0.00001).

**Figure 1:**
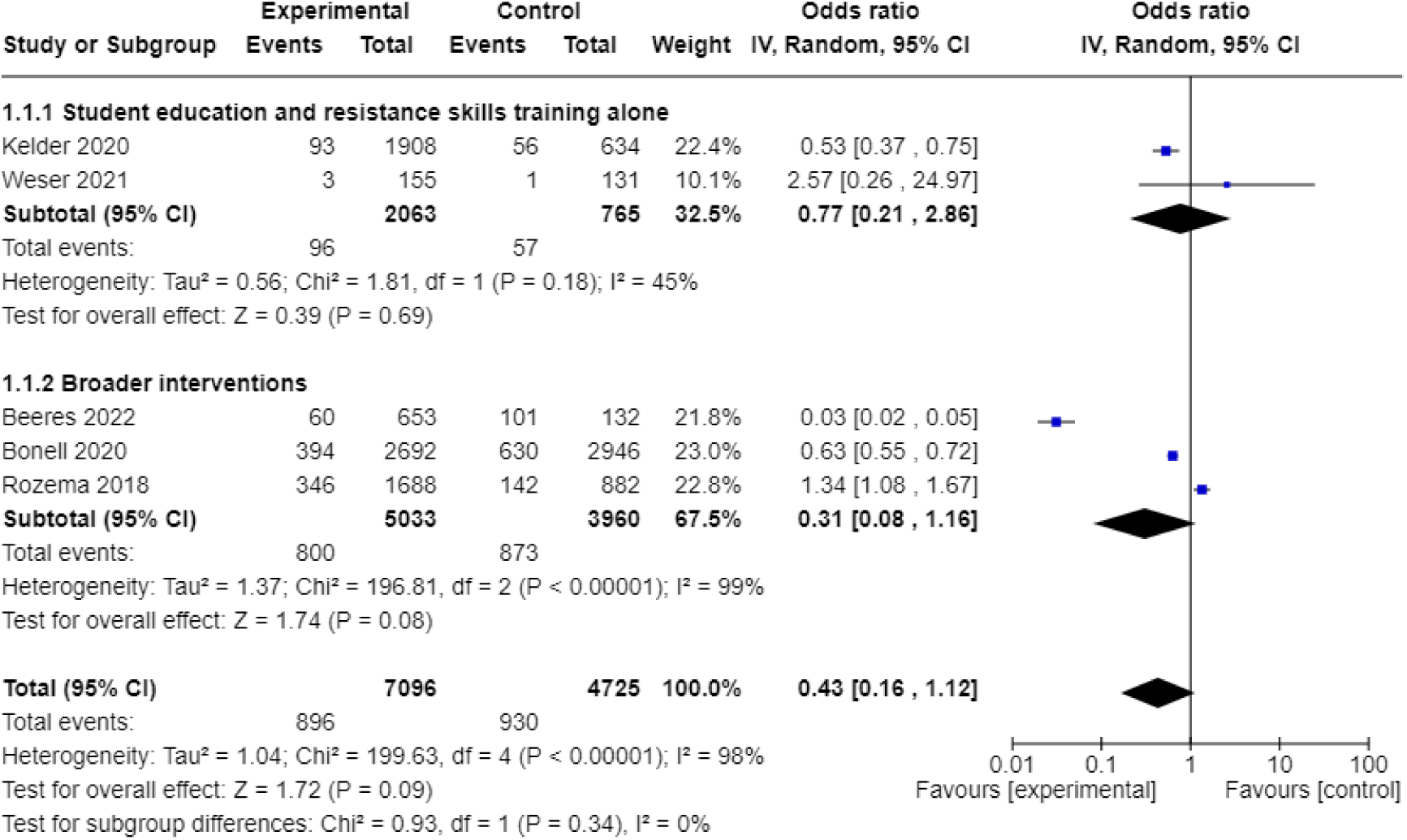
The prevention of e-cigarette use at longest follow-up.

#### Tobacco use

Pooled analysis of three studies showed that overall, school-based interventions were associated with a significant reduction in past 30-day tobacco use at longest follow-up (OR=0.59, 95% CI=0.39, 0.89, p=0.01; see Figure 2). However, there was substantial and significant heterogeneity between trials (I²=79%, p=0.009). Intervention and control groups did not significantly differ in terms of lifetime tobacco use (OR=1.01, 95% CI=0.65, 1.59, p=0.95), with considerable inconsistency between the two studies (I²=86%, p=0.008; Figure 3).

**Figure 2:**
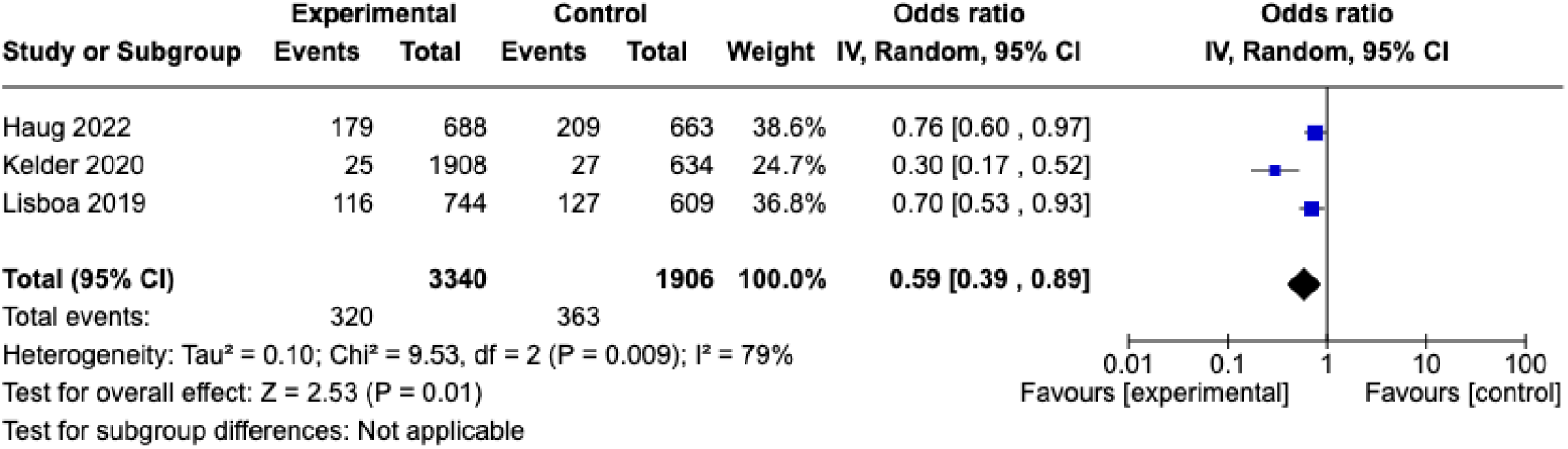
Past 30-day tobacco use at longest follow-up.

**Figure 3:**
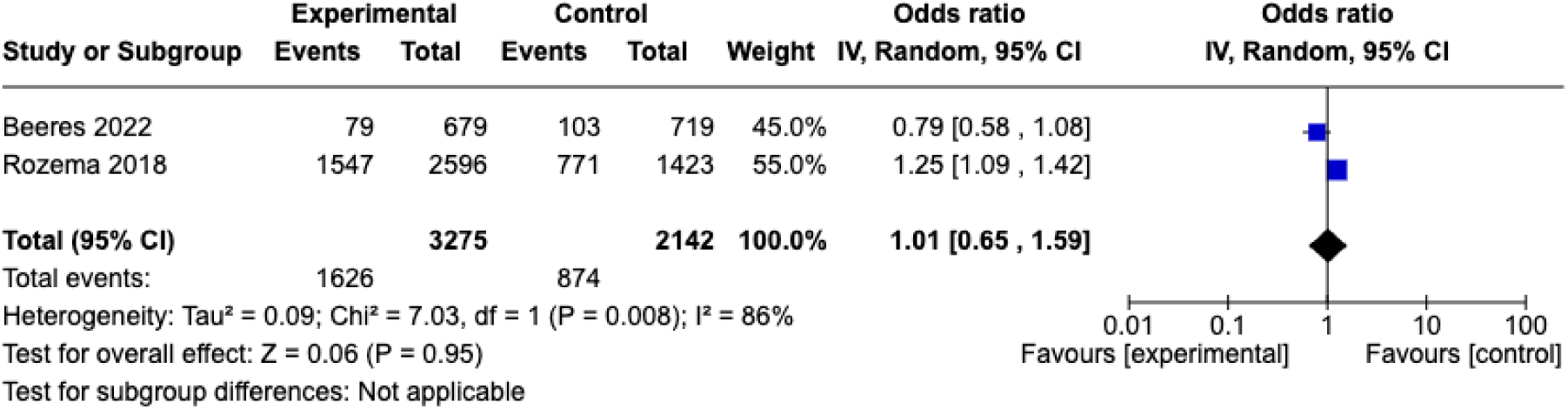
Lifetime tobacco use at longest follow-up.

#### Knowledge

School-based interventions had a small, yet significant effect on improving knowledge about e-cigarettes compared to controls (SMD=-0.38, 95% CI=-0.68, -0.08, p=0.01; Figure 4). However, there was considerable heterogeneity between studies (I^2^=78%, p=0.004).

**Figure 4:**
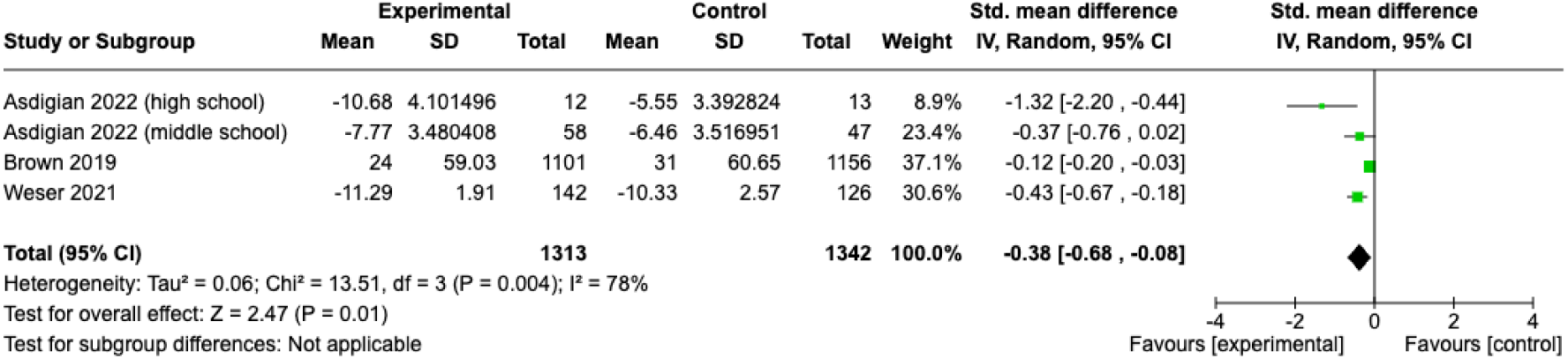
Knowledge about e-cigarettes at the first post-intervention timepoint.

#### Intentions to use e-cigarettes/tobacco

Overall, school-based interventions were associated with a significant reduction in students’ intentions to use e-cigarettes/tobacco (SMD=-0.15, 95% CI=-0.22, -0.07, p=0.0001), with no heterogeneity between studies (I^2^=0%; p=0.40; Figure 5).

**Figure 5:**
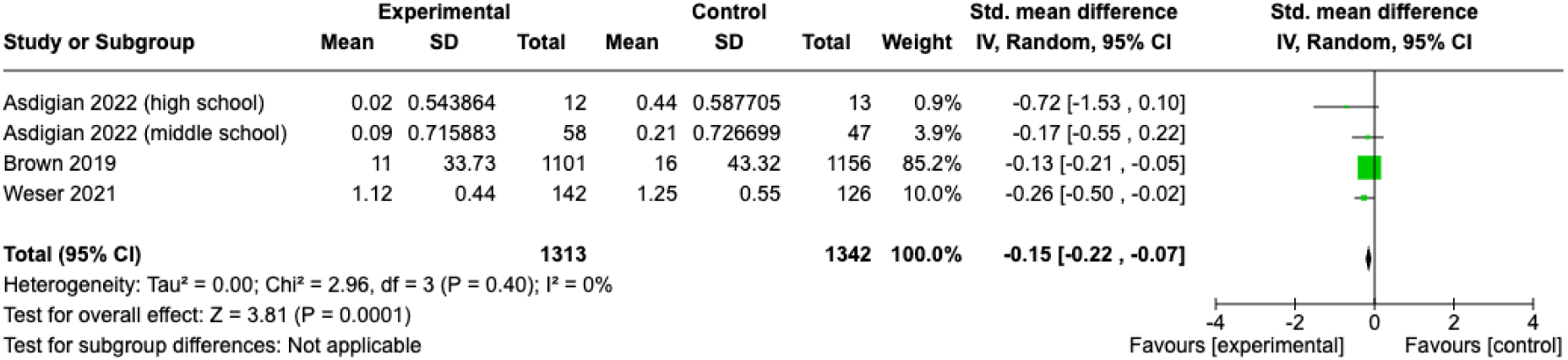
Intentions to use e-cigarettes at the first post-intervention timepoint.

#### Risky attitudes

Pooled analysis found a small, significant effect of school-based interventions on reducing risky attitudes towards e-cigarette use (SMD=-0.14, 95% CI=-0.22, -0.06; p=0.0007), with little variability between the three studies (I^2^=0%, p=0.39; Figure 6).

**Figure 6:**
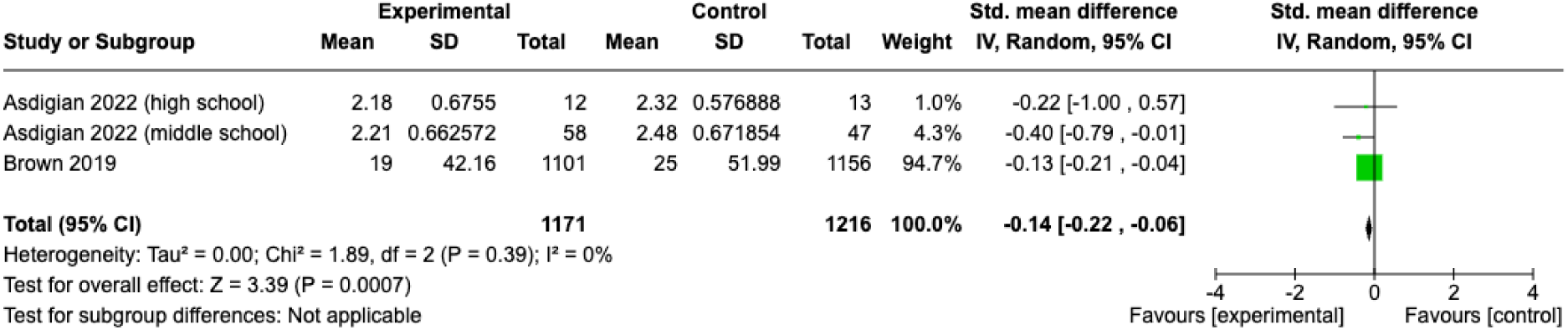
Risky attitudes towards e-cigarettes at the first post-intervention timepoint.

#### Harm perceptions

Overall, school-based interventions and controls groups did not significantly differ in terms of perceptions about the harms of e-cigarettes (SMD=-0.29, 95% CI=-0.73, 0.15, p=0.20; Figure 7). There was substantial and significant heterogeneity between trials (I^2^=69%, p=0.04).

**Figure 7:**
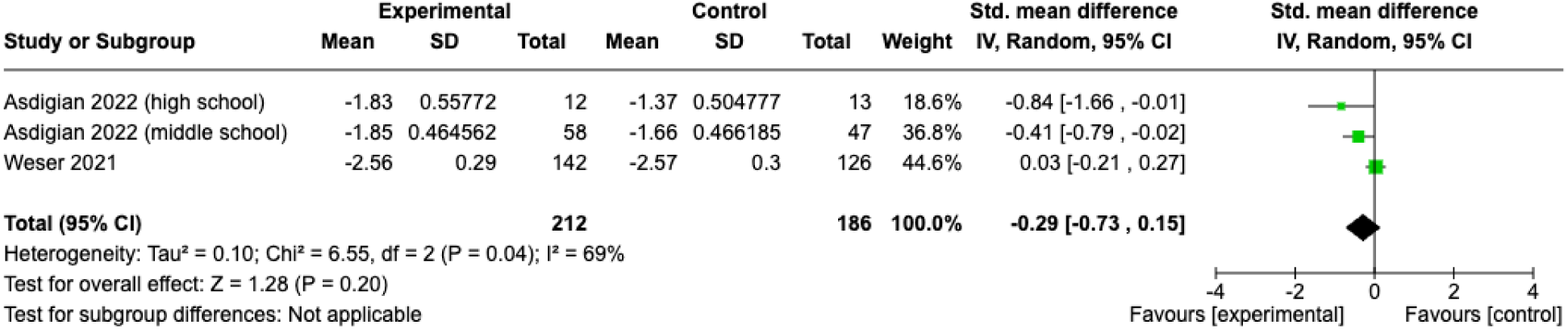
Harm perceptions at the first post-intervention timepoint.

### Risk of Bias and Quality of Evidence

Risk of bias assessments are provided in Supplemental Figures S1 and S2. Among randomised studies, one^38^ was judged at high risk of bias for past 30-day tobacco use due to missing outcome data, and there were some concerns for the remaining studies. This was primarily due to insufficient availability of information to determine whether there were any biases in outcome measurement or deviations from the intended intervention, along with missing outcome data. Among non-randomised studies, one^34^ was judged at serious risk of bias across outcomes due to bias in the classification of interventions and outcome measurement. There was also a moderate risk of bias judgement for all other studies, primarily due to potential bias in the measurement of outcomes, again related to insufficient availability of a prespecified protocol, and confounding.

Overall, we judged the quality of evidence to be low to moderate (see summary table in Supplemental Table S2). The quality of evidence for the prevention of e-cigarette use (both classroom education and skills training programs alone, as well as broader interventions), lifetime tobacco use, attitudes, and intentions was moderate. We judged the evidence related to past 30-day tobacco use, knowledge, and harm perceptions to be of low quality, primarily due to risk of bias and inconsistency between studies.

## Discussion

This is the first systematic review and meta-analysis to examine the efficacy of school-based preventive interventions for e-cigarette use. Our narrative synthesis showed that three^35^ ^36^ ^39^ of the six trials that reported specifically on the prevention of e-cigarette use found significant intervention effects in the expected direction, while one intervention inadvertently increased e-cigarette use.^44^ Our meta-analysis of five of these studies found that overall, school-based interventions were not significantly associated with the prevention of e-cigarette use. When considering tobacco use more broadly, which encompassed e-cigarettes in some studies, our narrative synthesis found that five trials^35^ ^36^ ^39^ ^40^ ^43^ reported significant intervention effects in the expected direction, whereas one increased tobacco smoking onset.^41^ Overall, our meta-analysis showed that school-based interventions were associated with a significant reduction in past 30-day tobacco use, albeit with substantial and significant heterogeneity between studies, but not lifetime tobacco use. Our narrative synthesis also revealed intervention effects on knowledge,^34^ ^37^ ^39^ ^42^ attitudes,^34^ ^37^ ^39^ normative beliefs,^34^ harm perceptions,^42^ future intentions to use tobacco,^37^ mental health outcomes,^36^ ^38^ and other substance use,^36^ ^38^ ^43^ with our meta-analyses finding that overall, school-based interventions were significantly associated with improvements in knowledge, intentions and attitudes at the first post-intervention timepoint. We judged the certainty in the body of evidence to be low-to-moderate overall.

The meta-analysis finding that school-based interventions were not significantly associated with the prevention of e-cigarette use highlights the need for more high-quality research to develop effective prevention programs, particularly given the existing evidence for school-based programs in preventing tobacco, alcohol and other drug use.^22^ ^23^ ^45^ When considering the findings of the narrative synthesis, although the components of the three interventions that prevented e-cigarette use varied, they all included a classroom education and skills training component that was delivered over multiple sessions (ranging between 4x25min lessons to a 10hr learning curriculum). The provision of developmentally appropriate information along with resistance and social skills training over a series of structured sessions aligns with established recommendations for effective school-based prevention of tobacco, alcohol and other drugs.^46^ Additionally, although the ‘CATCH My Breath’ program^39^ involved peer facilitators, all three programs kept teachers who received training within a central role; another characteristic previously linked to effective school-based tobacco and other drug prevention programs.^21^ ^46^ Two interventions^35^ ^36^ also included broader components, such as parental/adult involvement. For example, the core component of the ‘Tobacco-Free Duo’ intervention^35^ is a formal agreement between the student and a parent/adult to be tobacco-free for 3-years. A previous review of parent-based interventions found mixed results for the prevention of adolescent tobacco and alcohol use, with some interventions producing iatrogenic effects.^47^ However, the parent intervention components were more intensive than those in the present review and were typically not delivered as part of a broader school-based intervention.

The three interventions identified through the narrative synthesis as being efficacious in preventing e-cigarette use^35^ ^36^ ^39^ simultaneously produced improvements related to tobacco use, as did two additional interventions which included e-cigarettes within their broad definition of tobacco.^40^ ^43^ Although both additional interventions also involved classroom education along with resistance and social skills training, only the ‘HoLJouna Pono Drug Prevention Curriculum’^43^ was delivered over multiple sessions. In contrast, the ‘Education Against Tobacco (EAT)’ intervention^40^ was delivered as a one-off module via medical students; however, the total intervention dose (90mins) was similar to the efficacious ‘CATCH my breath’ program^39^ (100mins), and medical students may be considered akin to trained teachers. When tobacco outcomes were meta-analysed, school-based interventions were only associated with improvements in past 30-day tobacco use, but not lifetime use.

This likely relates to the small number of studies eligible for inclusion in the meta-analysis. Moreover, given previous reviews have demonstrated that school-based interventions can prevent tobacco use,^21^ this may also suggest that current interventions targeting both e-cigarettes and tobacco need to be improved upon. This could be done by ensuring alignment with the principles of effective school-based substance use prevention,^46^ or by improving engagement and implementation fidelity, which few studies reported on.

Notably, two interventions were associated with iatrogenic effects on the prevention of e-cigarette use^44^ and tobacco smoking onset.^41^ This included a “theme week”^44^ and school-wide policy which also produced null effects on e-cigarette use.^41^ From the limited descriptions, both interventions appeared broad, without specific classroom education and skills training, nor a theoretical underpinning. While the stated theories behind the interventions that successfully improved e-cigarette/tobacco use varied between the social cognitive theory, the theory of planned behaviour, and the theory of human functioning and school organisation, many of the interventions’ components aligned with the principles of a social competence and influence approach. This approach has previously exhibited the strongest evidence of effectiveness for school-based tobacco and other drug prevention.^21^ ^24^ ^46^ These findings highlight the critical importance of ensuring schools implement evidence-based programs to prevent e-cigarette/tobacco use.

Despite the lack of overall evidence generated for the prevention of e-cigarette and tobacco use, the meta-analyses indicated that school-based interventions were associated with significant improvements in knowledge, intentions, and attitudes. These constructs are theorised precursors of behaviour change,^29^ ^30^ yet, in the two studies that examined these constructs along with actual e-cigarette or tobacco use, only one^39^ observed positive intervention effects on all outcomes, which was at 16-month post-intervention timepoint. The other intervention, ‘Invite Only VR’,^42^ was associated with improved knowledge, but did not have a significant effect on e-cigarette use. This may have been due to the follow-up period only extending to 6-months, as a previous review found the effects of tobacco preventive interventions tend to only emerge after 1 year.^21^ Nevertheless, the four interventions identified via the narrative synthesis as having positive effects across knowledge,^34^ ^37^ ^39^ ^42^ attitudes,^34^ ^37^ ^39^ normative beliefs,^34^ harm perceptions,^42^ and future intentions to use tobacco^37^ all involved classroom education and skills training that was grounded in theory, again highlighting the potential of this intervention strategy. Three of the interventions^34^ ^37^ ^39^ also involved peer-led components, which has previously been shown to be effective at addressing adolescent substance use when compared to adult-led education.^48^ The use of eHealth intervention components, either in hybrid^34^ ^39^ or eHealth-only^42^ format, with delivery over multiple sessions also appeared beneficial. Previous research has found school-based eHealth interventions can be effective to prevent substance use, potentially because interactive and pre-programmed content can improve intervention fidelity and enhance student engagement and accessibility.^49^

Finally, although there were insufficient data for meta-analyses, the narrative synthesis found some interventions were associated with improvements in mental health^36^ ^38^ and other substance use, including reduced alcohol^36^ ^38^ and illicit drug use.^36^ ^43^ This is promising, given the strong associations between mental health and adolescent substance use, including e-cigarette use.^2^ Indeed, two of these studies^36^ ^43^ reported simultaneous improvements in e-cigarette and/or tobacco use, both of which involved classroom education and skills training over multiple sessions. Although the ‘Ready4Life’ intervention evaluated in the other study^38^ did not improve tobacco use, this may have been due to the limited follow-up period of 6-months as well the mobile-app being delivered outside of the classroom, with only 25% of students in the intervention condition engaging with the tobacco/e-cigarette smoking coaching topic.

## Limitations & Future Directions

Despite the rigorous methodology, this study had several limitations, including the small number of studies and significant heterogeneity present in many of our meta-analyses.

The quality of evidence was also low for some outcomes and effects were generally small and short-lived. In future, prospective registration of trial protocols would be beneficial to reduce potential bias. Only six out of the 11 identified studies measured actual e-cigarette and/or tobacco cigarette use, and there was substantial variation in the definitions across studies, making it difficult to tease out effects specifically on e-cigarettes compared to tobacco cigarettes and other tobacco products. The timeframes for the longest follow-up were also highly variable, ranging between 6-^42^ and 36-months.^36^ Longer-term follow-up periods are important when evaluating substance use preventive interventions, as effects are most likely to emerge in later adolescence as exposure to substances increases.^16^ ^21^ The remainder of studies focused on theoretical constructs related to e-cigarette/tobacco use, however, to fully understand the efficacy of such interventions in preventing e-cigarette use, assessment of actual substance use is required.

## Conclusions

Although some studies demonstrated that school-based e-cigarette preventive interventions can produce positive effects, some interventions negatively impacted students, highlighting the critical importance of supporting schools to identify and adopt evidence-based programs. Overall, our meta-analyses on a small subsample of studies found that school-based interventions were not effective in preventing e-cigarette or tobacco use, however, were associated with significant reductions in past 30-day tobacco use, which encompassed e-cigarettes in some studies. School-based interventions were also associated with improvements in knowledge, intentions, and attitudes, with some individual studies reporting improvements in mental health outcomes and other substance use. Classroom education and student skills training that is grounded in theory and delivered by trained teachers was commonly associated with beneficial program effects. Peer-led and eHealth components also appeared promising to improve precursors of behaviour change, however more research, over the longer-term, is required to evaluate whether this translates into effects on e-cigarette use. Given previous research has demonstrated that school-based interventions can be effective in preventing tobacco, alcohol and other drug use, school-based interventions may have unmet potential for addressing e-cigarette use and more high-quality research is required to develop efficacious interventions.

## Data Availability

All data produced in the present study are available upon reasonable request to the authors

---

**Table S1.**
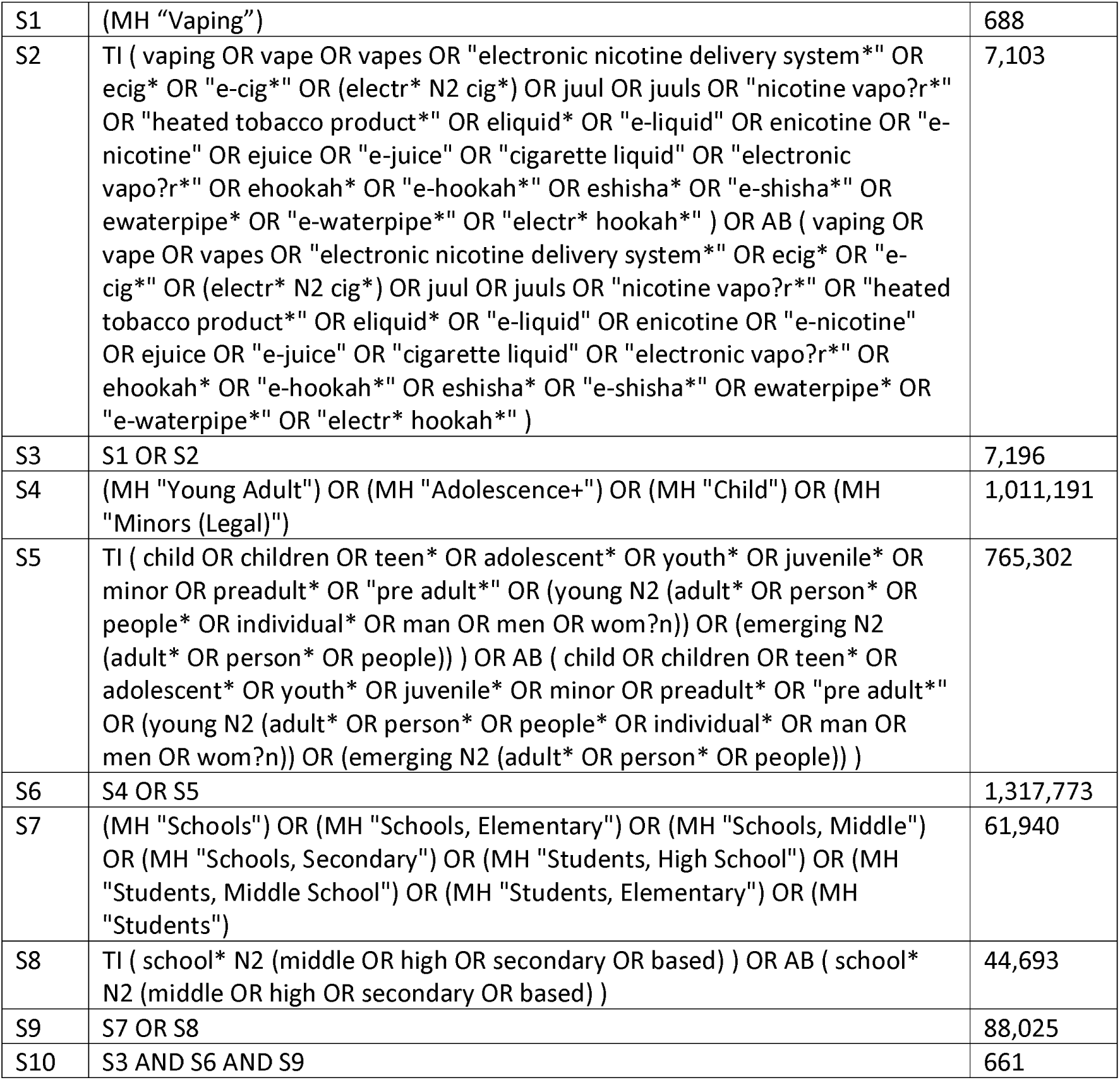
Example Search Strategy (CINAHL)

|  |  |  |
| --- | --- | --- |
| S1 | (MH "Vaping") | 688 |
| S2 | TI ( vaping OR vape OR vapes OR "electronic nicotine delivery system*" OR ecig* OR "e-cig*" OR (electr* N2 cig*) OR juul OR juuls OR "nicotine vapo?r*" OR "heated tobacco product*" OR eliquid* OR "e-liquid" OR enicotine OR "e-nicotine" OR ejuice OR "e-juice" OR "cigarette liquid" OR "electronic vapo?r*" OR ehookah* OR "e-hookah*" OR eshisha* OR "e-shisha*" OR ewaterpipe* OR "e-waterpipe*" OR "electr* hookah*" ) OR AB ( vaping OR vape OR vapes OR "electronic nicotine delivery system*" OR ecig* OR "e-cig*" OR (electr* N2 cig*) OR juul OR juuls OR "nicotine vapo?r*" OR "heated tobacco product*" OR eliquid* OR "e-liquid" OR enicotine OR "e-nicotine" OR ejuice OR "e-juice" OR "cigarette liquid" OR "electronic vapo?r*" OR ehookah* OR "e-hookah*" OR eshisha* OR "e-shisha*" OR ewaterpipe* OR "e-waterpipe*" OR "electr* hookah*" ) | 7,103 |
| S3 | S1 OR S2 | 7,196 |
| S4 | (MH "Young Adult") OR (MH "Adolescence+") OR (MH "Child") OR (MH "Minors (Legal)") | 1,011,191 |
| S5 | TI ( child OR children OR teen* OR adolescent* OR youth* OR juvenile* OR minor OR preadult* OR "pre adult*" OR (young N2 (adult* OR person* OR people* OR individual* OR man OR men OR wom?n)) OR (emerging N2 (adult* OR person* OR people)) ) OR AB ( child OR children OR teen* OR adolescent* OR youth* OR juvenile* OR minor OR preadult* OR "pre adult*" OR (young N2 (adult* OR person* OR people* OR individual* OR man OR men OR wom?n)) OR (emerging N2 (adult* OR person* OR people)) ) | 765,302 |
| S6 | S4 OR S5 | 1,317,773 |
| S7 | (MH "Schools") OR (MH "Schools, Elementary") OR (MH "Schools, Middle") OR (MH "Schools, Secondary") OR (MH "Students, High School") OR (MH "Students, Middle School") OR (MH "Students, Elementary") OR (MH "Students") | 61,940 |
| S8 | TI ( school* N2 (middle OR high OR secondary OR based) ) OR AB ( school* N2 (middle OR high OR secondary OR based) ) | 44,693 |
| S9 | S7 OR S8 | 88,025 |
| S10 | S3 AND S6 AND S9 | 661 |

**Figure S1.**
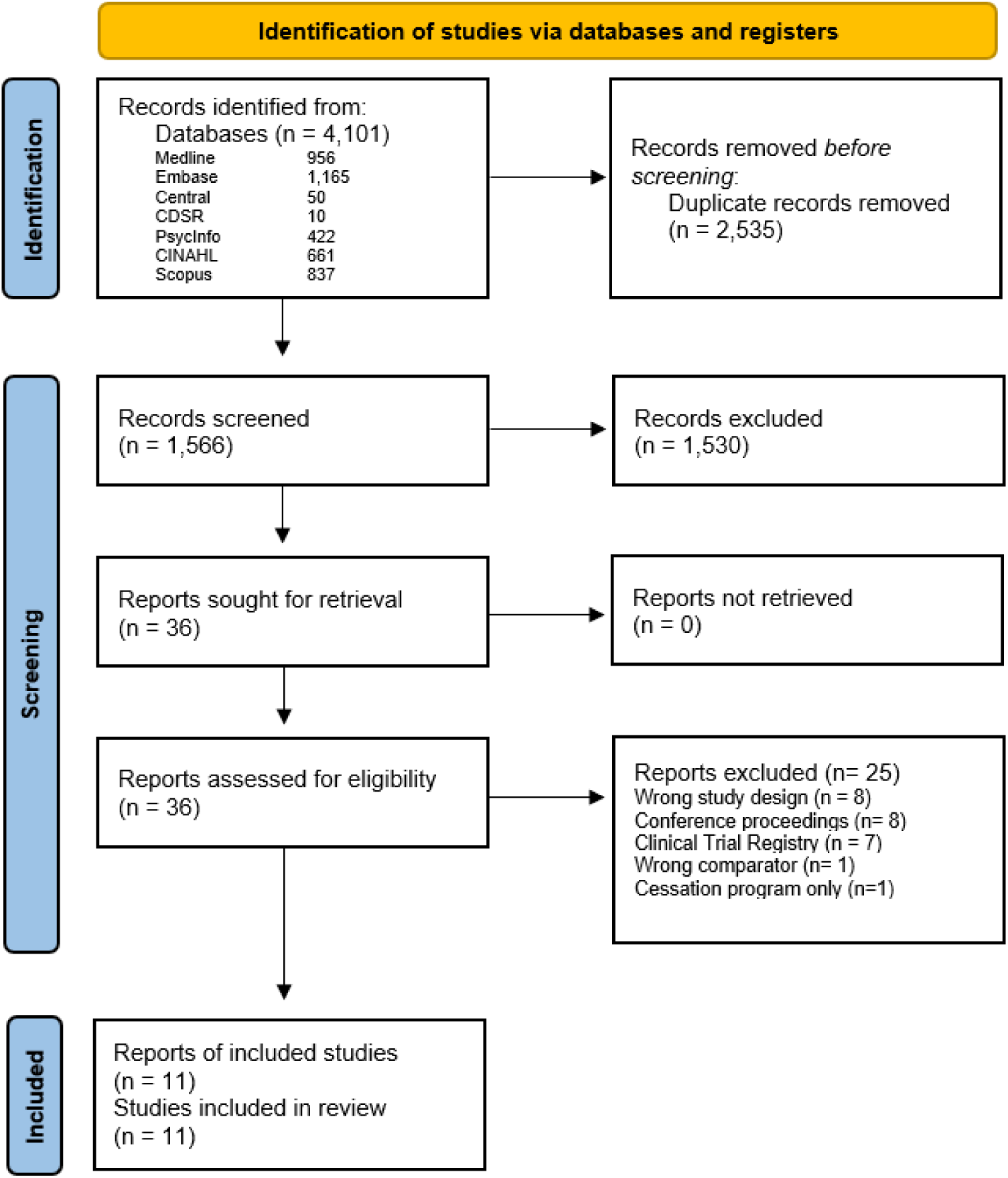
PRISMA flow diagram.

**Figure S2.**
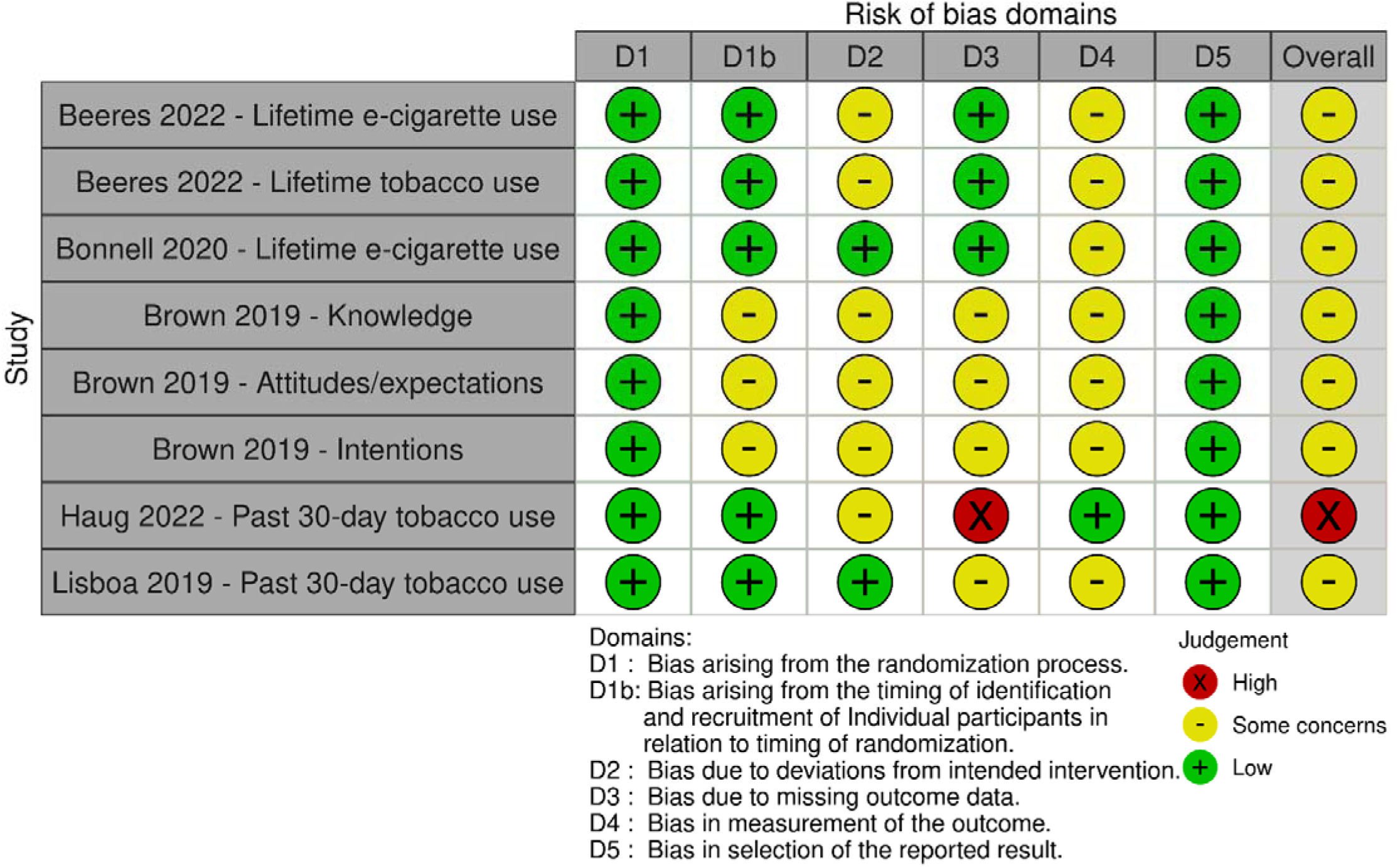
Risk of Bias of Randomised Studies.

**Figure S3.**
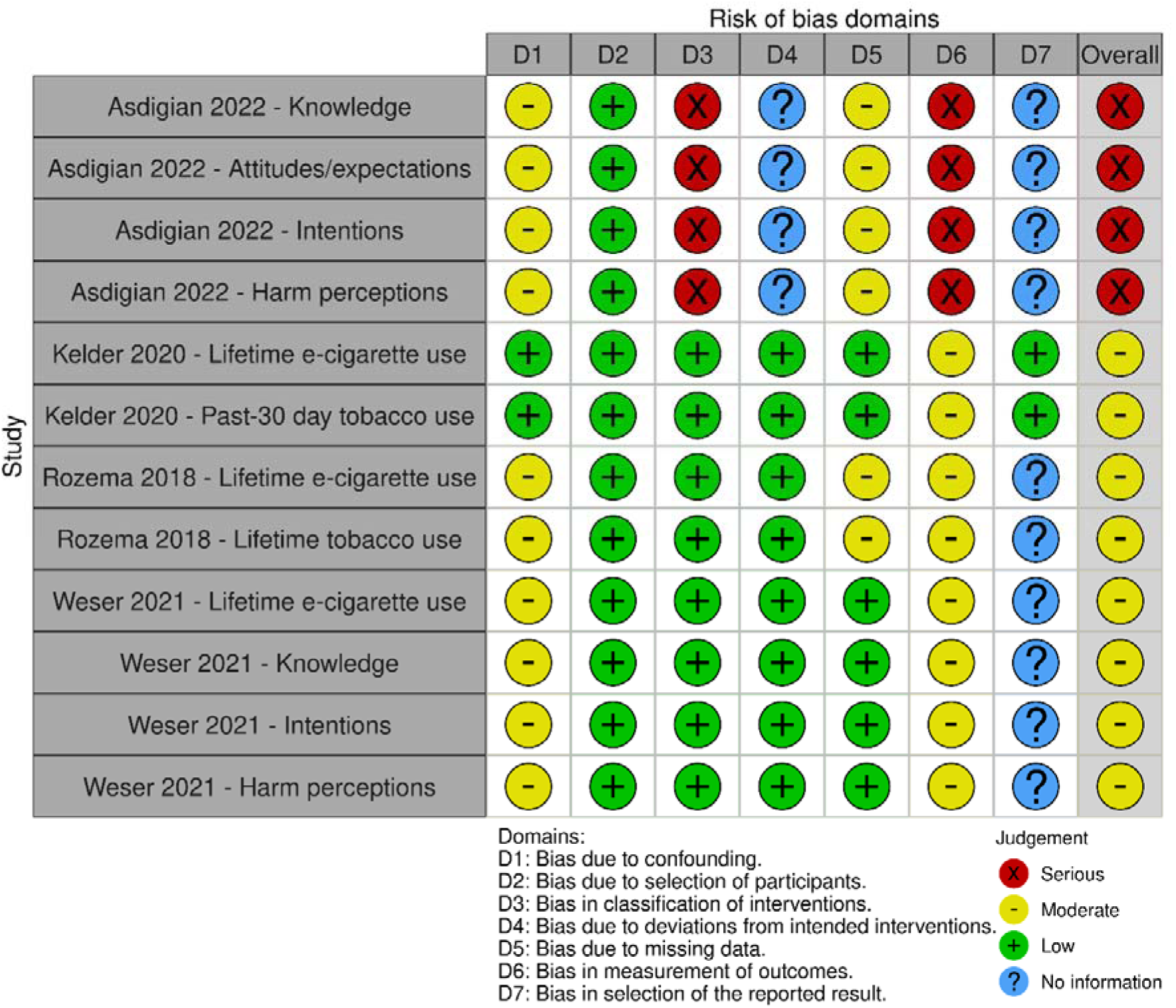
Risk of Bias of Non-Randomised Studies.

**Table S2.**
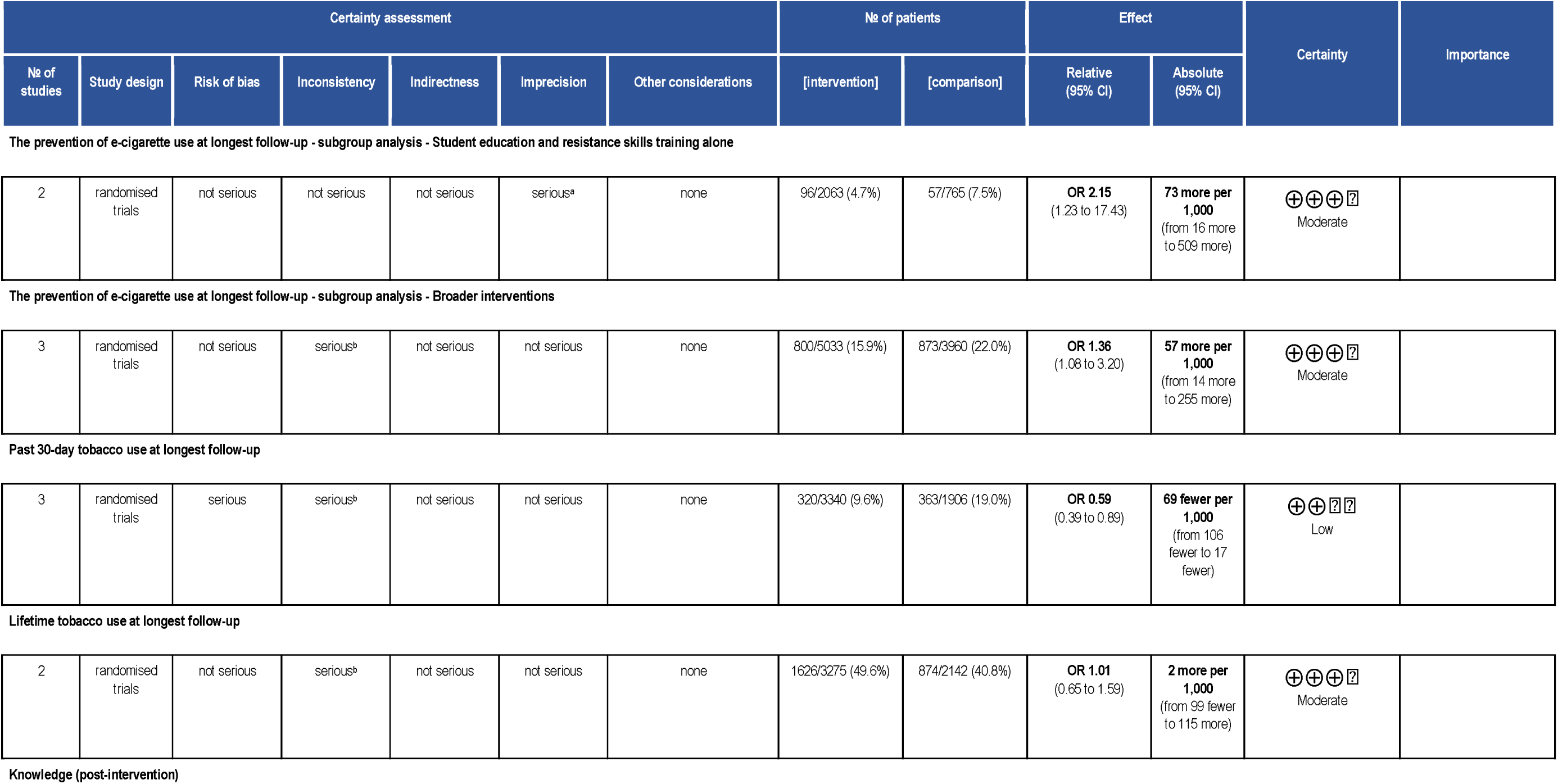
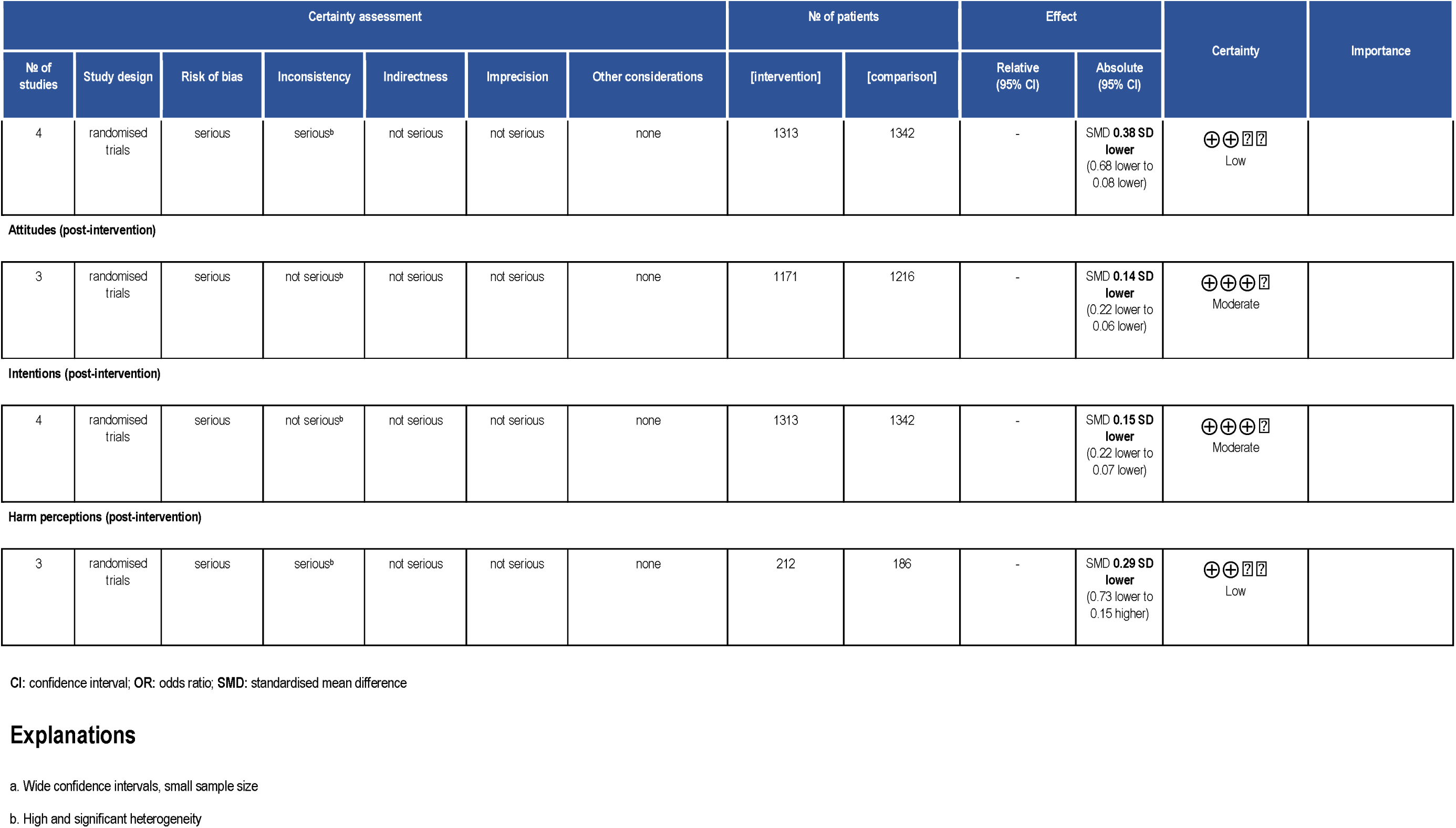
Summary of Findings Table (Quality of Evidence Ratings)

